# Inflammation-based risk score predicts kidney survival in children and adults with chronic kidney disease

**DOI:** 10.64898/2026.07.01.26357013

**Authors:** Hendrik Bartolomaeus, Rosa Reitmeir, Jakob Versnjak, Jonas Hofstetter, Felix Behrens, Alex Yarritu, Paul M. Bonnekoh, Max C. Liebau, Aysun K. Bayazit, Ali Duzova, Nur Canpolat, Ipek Kaplan Bulut, Karolis Azukaitis, Lukasz Obrycki, Nicola Wilck, Marcus Weitz, Alma Zernecke, Anette Melk, Uwe Querfeld, Marcus Kelm, 4C Study Consortium, Franz Schaefer, Johannes Holle

## Abstract

Chronic kidney disease (CKD) is accompanied by systemic inflammation, but whether inflammatory proteins improve risk stratification beyond kidney function and albuminuria remains unclear. We profiled baseline serum samples from 683 children with CKD in the prospective European 4C study using the Olink Target 96 Inflammation panel and related protein levels to kidney outcomes over 8 years. eGFR and albuminuria were the dominant determinants of the inflammation-related serum proteome. Nonetheless, a four-protein inflammation score (iScore; CD40, CD137, PD-L1 and CX3CL1), defined from protein residuals independent of eGFR and albuminuria, stratified kidney survival and predicted CKD progression beyond established clinical risk factors (adjusted hazard ratio per elevated protein, 1.11; 95% CI, 1.01–1.22). The score showed concordant associations in 2,770 UK Biobank participants with reduced eGFR (adjusted hazard ratio, 1.08; 95% CI, 1.02–1.14). Kidney single-cell transcriptomic analysis mapped these axes to immune–parenchymal communication programs in CKD, supporting inflammation-based risk stratification across the life course.

## Introduction

Chronic kidney disease (CKD) is a highly relevant, global health concern, with a current prevalence of around 14% of the adult population^1^. According to recent projections, CKD will become the fifth leading cause of death worldwide by 2040^2^. Once established, CKD typically follows a progressive course, although the rate of decline is highly variable between individuals. Despite recent advances in medical care of CKD patients, including inhibitors of the renin angiotensin system (RASi)^3^, non-steroidal mineralocorticoid receptor (MR) antagonist^4^ and sodium glucose linked transporter 2 (SGLT2) inhibitors^5^, the risk remains substantial, and several drivers of CKD are so far unaddressed. Importantly, CKD is accompanied by low-grade chronic systemic inflammation (SCI), which significantly contributes to the multimorbidity and excessive mortality that is characteristic of CKD^6,7^. Experimental evidence supports SCI as a modifiable driver of CKD progression, with targeted anti-inflammatory interventions attenuating renal dysfunction and fibrosis in animal models of CKD^8-10^. However, to develop effective therapeutic strategies for clinical use, a comprehensive analysis of SCI and its impact on disease progression in patients with CKD is needed.

Of note, SCI is not a unique mechanism in CKD, but implicated across multiple chronic disease states^11^. CKD in adult patients is commonly a consequence of long-lasting exposition to comorbidities and external risk factors (e.g. diabetes, hypertension, life-style)^12^. This makes it very difficult to specifically characterize associations between CKD and SCI. In contrast, pediatric patients usually suffer from primary CKD, most frequently due to congenital anomalies of the kidney and urinary tract (CAKUT)^13^. We therefore see a unique benefit of exploring SCI-CKD associations in children in the absence of confounding comorbidities and validation of key results in the adult population in a rare-to-common approach.

Here, we present a first comprehensive analysis of SCI in CKD and its potential to predict kidney outcomes using a targeted proteomics approach. We therefore analysed samples from the Cardiovascular Comorbidities in Children with CKD (4C) study^14^, one of the largest pediatric CKD cohorts, and validated our findings in participants of the UK Biobank (UKBB)^15^.

## Methods

### 4C Cohort

The 4C Study, initiated in 2009, prospectively followed 704 patients aged 6 to 17 years with a baseline estimated glomerular filtration rate (eGFR) of 10 to 60 mL/min/1.73m^2^ in 55 pediatric nephrology units across 12 European countries^14^. The 4C Study was approved by the Ethics Committee of Heidelberg University (S-032/2009) and the institutional review boards at each participating institution. Written informed consent was obtained from all legal guardians and participants as appropriate. The study is registered at ClinicalTrials.gov (NCT01046448).

Kidney function was assessed by the eGFR as described previously^16^. Albuminuria was quantified using albumin/creatinine ratio (UACR). For numeric variables, when given, standard deviation scores (sds) based on age and sex were considered to account for the known variability in pediatric cohorts. Due to the large range of UACR, the values were log-transformed. All clinical variables including units used within this analysis are listed in Supplemental Methods.

We used a combined kidney endpoint (CKE) for CKD progression composed of eGFR loss of more than 50% since study entry and/or start of kidney replacement therapy including dialysis (hemo and peritoneal) or transplantation (preemptive and non-preemptive). Additionally, death was also included in this composite endpoint.

### OLINK proteomics

At study entry, systemic inflammation was assessed using a targeted serum proteomics approach (OLINK Target 96 Inflammation, Olink Proteomics, Uppsala, Sweden), measuring 92 serum proteins. A total of 683 samples were analysed. We evaluated potential sample differences across assay plates, repeated or not repeated sample measurements, and quality control flags (pass or warning). No systematic differences were observed, so all samples were used for subsequent analyses. Seventy seven out of 92 serum proteins were selected for further analyses, fulfilling the criterion of reported values above the limit of detection (LOD) in at least 70% of the samples. Values below LOD were imputed using the log-transformed square root of the exponentiated maximum LOD. For seven proteins with a missing rate between 70% and 90%, we evaluated whether their absence was random or associated with kidney function; no significant differences were observed across CKD stages, accordingly they were not considered for further analyses (see Figure S1).

### Statistical analyses

All statistical analyses were performed in R (version 4.5.0). Group comparisons were conducted using Wilcoxon rank-sum tests or Kruskal-Wallis tests for continuous variables and χ^2^ tests for categorical variables. Relationships between continuous variables were assessed by Spearman’s rank correlation. Associations between the Olink proteome and clinical parameters were investigated using multivariable linear and Cox proportional hazards models with adjustment for relevant covariates. Potential confounding effects were assessed using *metadeconfoundR* (version 1.0.2). In addition, random forest models with k-fold cross validation were constructed to determine the predictive power of protein and clinical features. For multiple testing scenarios, the Benjamini-Hochberg procedure was used to control the false discovery rate (FDR) at 5%. Statistical significance was defined as an FDR adjusted p-value of < 0.05.

Detailed descriptions of data preprocessing, variable definitions, and model constructions are provided in the supplement. All scripts are publicly available on GitHub (https://github.com/rosareitmeir/4C_OLINK_CKDprogression). For exploration of individual proteins across major clinical variables we provide an interactive web application with spearman correlations for continuous variables and Kruskal-Wallis test followed by Mann-Whitney U-test with Benjamini-Hochberg false discovery rate correction post-hoc (https://hbartolomaeus.shinyapps.io/olink_inflammation_4c/).

## Results

### Chronic kidney disease shapes inflammation-related serum proteome

We analysed stored biomaterial from the pan-European 4C Study, a longitudinal cohort of children with CKD stages 3–5 not requiring kidney replacement therapy at study entry. A total of 683 children with available baseline serum samples were included (mean age, 12.1 ± 3.3 years; 34.7% female), with a mean baseline eGFR of 28.8 ± 11.4 mL/min/1.73 m². CAKUT was the most common underlying kidney disease (68.8%), and eGFR was similarly distributed across diagnostic categories. Participants were recruited across twelve European countries, with the largest subgroup enrolled in Turkey (Fig. S2).

Using the OLINK Target 96 Inflammation panel, we quantified inflammation-related serum proteins at baseline. After standard quality control, to estimated the variance explained by different metadata categories we used LASSO regression models fitted separately for each protein. CKD-related variables (e.g., eGFR and UACR) explained the largest proportion of variance across most detected proteins (Fig. 1A). This was confirmed by principal component analysis (PCA), in which both PC1 (27% variance) and PC2 (11%) were associated with eGFR and UACR (Fig 1A-C). Consistent with these findings, multivariable linear modeling identified signficiant negative associations of 46 of 77 proteins with eGFR and positive associations of 59 proteins with UACR (Fig. S3). After adjustment for age, sex, BMI, country, underlying kidney disease, and mutual adjustment for eGFR and UACR, 41 and 33 associations remained significant, respectively (Fig. 1D). Associations of proteins with eGFR and UACR were independent of protein size and charge (Fig S4).

**Figure 1.**
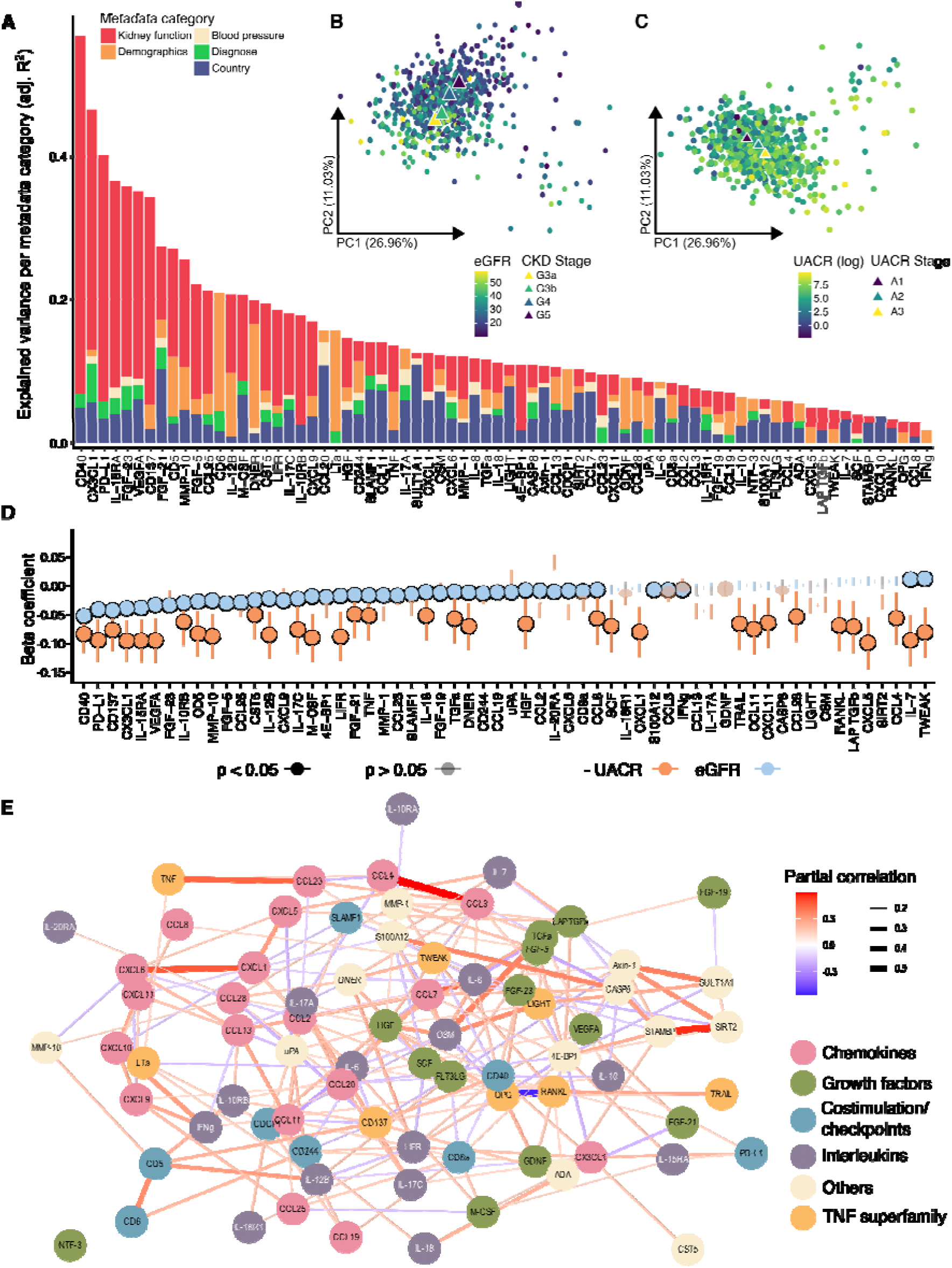
Kidney function is the major determinant of the inflammation-related plasma proteome in pediatric CKD. (A) Variance in individual inflammation-related plasma proteins explained by different metadata categories, quantified using LASSO regression models fitted separately for each protein and grouped by category. Bars are ordered by total explained variance. (B, C) Principal component analysis (PCA) of inflammation-related plasma proteins, colored by baseline eGFR and annotated by CKD stage (B) or colored by urinary albumin-to-creatinine ratio (UACR) and annotated by albuminuria stage (C). (D) Associations of individual proteins with baseline eGFR and UACR in multivariable linear regression models. Points indicate standardized regression estimates and error bars indicate 95% confidence intervals after adjustment for age, sex, BMI, country, underlying kidney disease, and mutual adjustment for eGFR and UACR. Orange points indicate associations with UACR and blue points indicate associations with eGFR. Shaded points and error bars denote non-significant associations when controlling the false discovery rate across all tests at 5%. (E) Network analysis was performed using partial correlation, adjusted for kidney function and UACR, across all proteins. Proteins were colored according to its biological function.

By contrast, demographic variables explained substantially less variance overall, although they accounted for the largest share of variance in a small subset of proteins, including for example CD6 and DNER (Fig. 1A). Country and underlying kidney disease showed smaller but detectable effects, consistent with the PCA (Fig. S5). To further quantify the impact of underlying kidney diseae and country of recruitment, we performed contrast analyses using CAKUT and Turkey as the respective reference groups. Glomerulopathies differed most strongly from CAKUT, with 12 proteins increased and 4 decreased, including prominently higher CX3CL1 and FGF-23 levels (Fig. S6). Across recruiting countries, several proteins differed in abundance, with generally lower levels observed at smaller study sites compared with Turkey, potentially reflecting biological, environmental, or pre-analytical influences (Fig. S7).

To assess the structure of the inflammatory proteome beyond the dominant effects of kidney function, we generated a partial-correlation network adjusted for eGFR and UACR. This analysis revealed persistent inter-protein relationships, indicating that the proteomic signature retains structured covariance beyond shared variation explained by kidney function and UACR (Fig. 1E).

Together, these analyses identify kidney function as the major determinant of the inflammation-related serum proteome in pediatric CKD. To facilitate reuse of the dataset, we additionally provide an interactive web application for exploration of individual proteins across major clinical variables. (https://hbartolomaeus.shinyapps.io/olink_inflammation_4c/).

### Inflammatory serum proteins are associated with kidney outcome

We hypothesized that baseline inflammatory signatures are associated with subsequent kidney outcome. We therefore analysed the occurrence of the composite kidney endpoint (CKE; defined as eGFR loss of more than 50%, start of kidney replacement therapy, or death), over up to 96 months of follow-up. Considerable dropout was observed over time, most commonly due to transition to adult nephrology care (Fig. 2A).

**Figure 2.**
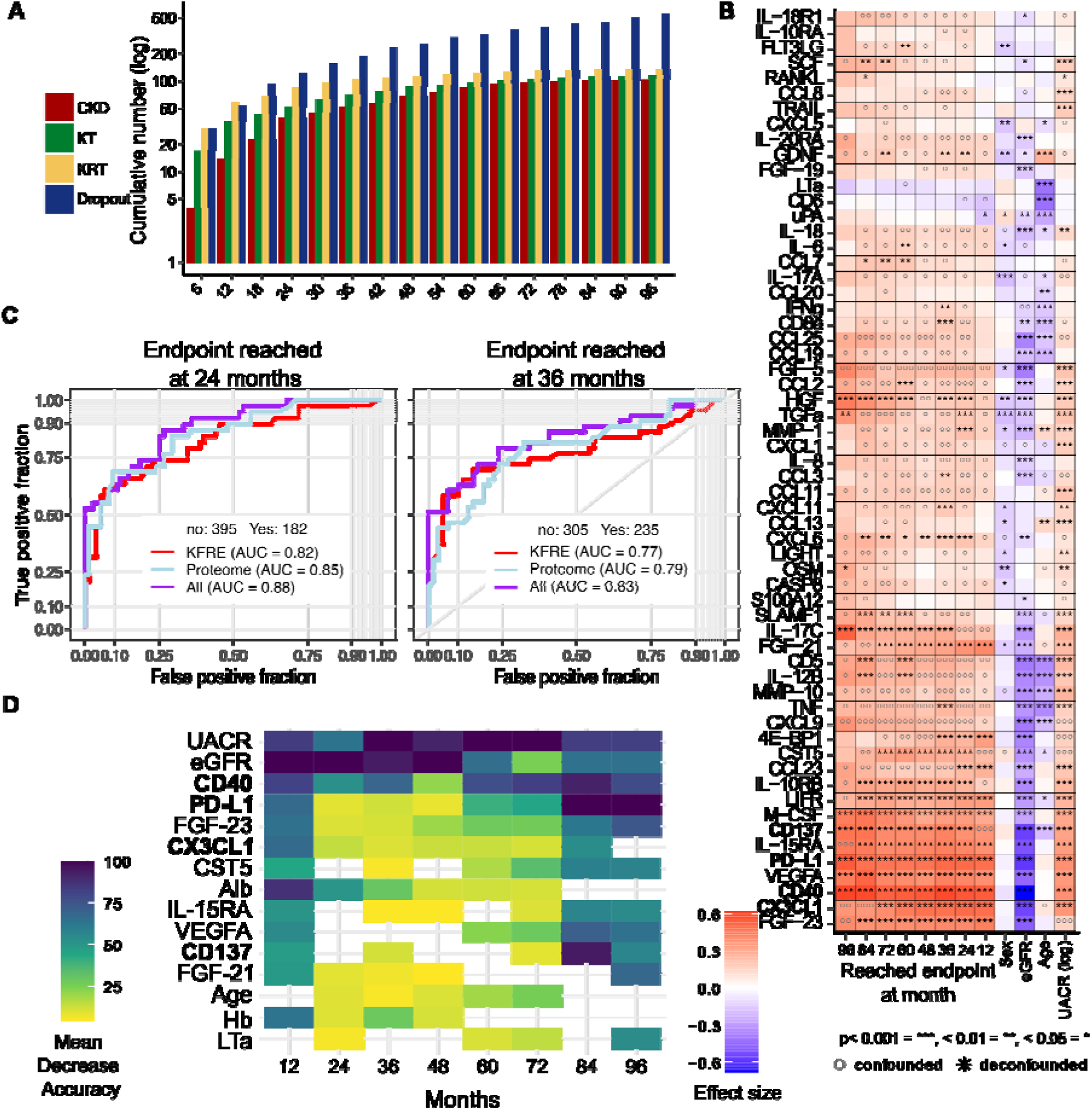
Baseline inflammatory plasma proteins predict kidney outcome in pediatric CKD. (A) Distribution of follow-up status across time points from baseline to 96 months. (B) Confounder-aware associations between baseline plasma proteins and the composite kidney endpoint (CKE) at discrete follow-up time points. Colors indicate effect size. Symbols denote whether associations remained significant after accounting for potential confounders, including eGFR, UACR, routine laboratory variables, demographic variables, and country, when controlling the false discovery rate across all tests at 5%. (C) Random forest models for prediction of the CKE at 24 and 36 months. Receiver operating characteristic curves are shown for models including KFRE variables alone, proteome variables alone, or all variables combined. Area under the curve (AUC) is indicated for each model. (D) Variable importance across combined random forest models for prediction of the CKE at different time points. Colors indicate mean decrease in accuracy, with higher values reflecting greater importance for model performance.

As a first step, we performed confounder-aware analyses to test associations between the baseline serum proteome and the CKE at discrete follow-up time points while accounting for potential confounders, including eGFR, UACR, routine laboratory values, demographic variables, and country (Fig. 2B; full heatmap in Fig. S8). This revealed a consistent protein signature associated with reaching the CKE from 12 to 96 months of follow-up. The majority of protein-endpoint associations were driven by changes in eGFR and UACR, emphasizing the important role of kidney function in determining inflammatory protein levels. However, several associations remained independent of kidney function. For example, IL-15RA, PD-L1, and FGF-23 were consistently positively associated with the CKE and were not explained by other risk factors at most time points. In contrast, associations of CXCL9 and TNF with the CKE were largely attributable to kidney function.

Current equations for the prediction of kidney failure, such as the Kidney Failure Risk Equation (KFRE), rely on age, sex, eGFR and UACR. Adding inflammatory protein information carries the potential to refine such predictions. We thus applied applied random forest models to assess the predictive performance of inflammatory proteins for the CKE (Fig. 2C; full summary for all endpoints in Table S1). Using an 80:20 training-test split, we first compared the performance of proteome-based models with models based on KFRE variables alone. We then tested whether addition of proteomic features to the full set of clinical variables improved prediction. Across combined models, we further examined which variables contributed most consistently to predictive performance across time points (Fig. 2D; full heatmap in Fig. S9). As expected, eGFR and UACR ranked among the most influential predictors. However, several inflammatory proteins, including CD40, FGF-23, and CX3CL1, also showed high importance for prediction of the CKE, consistent with the confounder-aware analyses.

Together we show that inflammatory serum proteins are associated with a CKE in pediatric CKD and improve prediction beyond eGFR and UACR as established clinical risk factors.

### Inflammation-based risk score improves patient risk stratification

To integrate inflammatory proteins with prognostic value into a single interpretable score, we prioritized proteins that showed consistent associations with the CKE in confounder-aware analyses and contributed to outcome prediction in random forest models. Several proteins emerged consistently across these analyses, including FGF-23, CD40, CD137, PD-L1, and CX3CL1. Because FGF-23 is heavily influenced by CKD-associated mineral and bone diseae^17^ and has already been linked to progression of CKD^18^, we focused on four less-established candidates, CD40, CD137, PD-L1, and CX3CL1, that represented complementary immune-inflammatory axes.

A local residual correlation network further showed that CD40, CD137, PD-L1, and CX3CL1 were embedded in partly distinct proteomic neighborhoods after adjustment for eGFR and UACR, supporting their non-redundant contribution to an integrated inflammatory score (Fig. S10). For each of the four proteins, elevated levels were defined as residual values in the highest 20% after adjustment for eGFR and UACR (Fig. 3A). Patients were then categorized according to an inflammation score (iScore), defined by the number of elevated proteins: i1, no elevated proteins; i2, one to two elevated proteins; and i3, three to four elevated proteins. By design, eGFR and UACR were similar across iScore categories (Fig. 3C), allowing assessment of inflammatory risk beyond conventional kidney function and UACR-based classification.

**Figure 3.**
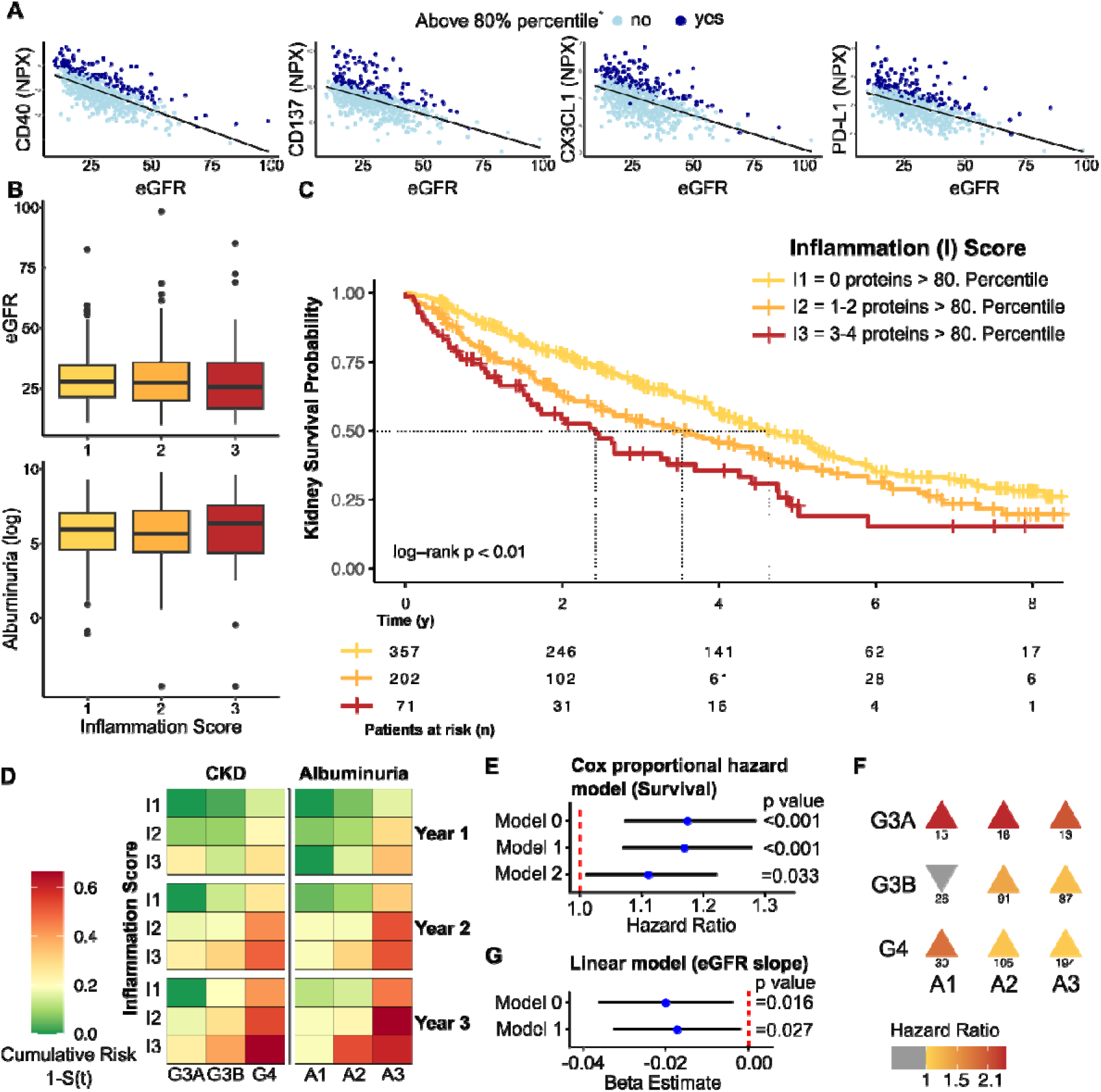
An inflammation-based risk score improves risk stratification for CKD progression in children. (A) Baseline levels of CD40, CD137, CX3CL1, and PD-L1 in relation to eGFR. Elevated protein levels were defined as residual values above the 80^th^ percentile after adjustment for eGFR and albuminuria. (B) Distribution of baseline eGFR and albuminuria across inflammation score (iScore) categories. Patients were grouped according to the number of elevated inflammatory proteins: I1, 0 proteins; I2, 1–2 proteins; I3, 3–4 proteins. (C) Kaplan–Meier analysis for the composite kidney endpoint according to iScore category. Tick marks indicate censoring, and numbers at risk are shown below the plot. P-value was calculated by log-rank test. (D) Heatmap showing cumulative risk of the composite kidney endpoint at years 1, 2, and 3 across CKD stage and albuminuria stage, further stratified by iScore category. (E) Association of iScore with the composite kidney endpoint in Cox proportional hazards models with increasing degrees of adjustment. Points indicate hazard ratios and horizontal lines indicate 95% confidence intervals. Model 0 only iScore, Model 1 with inclusion of age, sex, eGFR, albuminuria, and Model 2 with further adjustment for underlying kidney disease and laboratory values (full Models 1 and 2 in Figure S11). (F) Hazard ratios for the association of iScore with the composite kidney endpoint across strata of CKD stage and albuminuria. Color indicates hazard ratio and numbers indicate the number of participants per subgroup. (G) Association of iScore with relative eGFR change within the first year after study entry. Linear regression coefficients with 95% confidence intervals are shown for a model with only iScore (Model 0) and iScore with KFRE variables (Model 1, full models in Fig. S11).

Median kidney survival decreased stepwise across iScore categories, with the time to CKE in 50% of part shortened by nearly 50% in patients with three to four elevated proteins compared with patients without increased proteins (Fig. 3C). Across CKD stage and UACR strata, the cumulative risk of reaching the CKE within years 1–3 was further refined by iScore category (Fig. 3D; full description in Fig. S11). This association persisted in multivariable Cox regression adjusting for established clinical risk factors, including baseline eGFR, UACR, and additional clinical covariates (Fig. 3E; full models in Fig. S12). The iScore also stratified risk across nearly all eGFR and UACR stages represented in the 4C cohort (Fig. 3F).

Finally, we assessed short-term kidney function decline within the first year after study entry. Higher iScore was associated with greater relative eGFR loss, with similar effect estimates after adjustment for KFRE variables (Fig. 3G). Addition of the iScore significantly improved model fit compared with a model based on established clinical predictors alone. Together, these data show that an inflammation-based score refines risk prediction for CKD progression in children beyond established risk factors, despite the close relationship between inflammatory protein levels and kidney function.

### Validation of the inflammation-based risk score in the UK Biobank

We next evaluated the iScore in adult participants of the UK Biobank (UKBB; baseline characteristics in Table S2). Elevated levels of the corresponding proteins were defined analogously using the available proteomic data (Fig. S13). To approximate the CKD phenotype of the 4C cohort, survival analyses were restricted to participants with baseline eGFR <60 mL/min/1.73 m². In total, 2,770 individuals were included (62.3 ± 6.2 years; 42.4% female), with a mean eGFR of 51.18 ± 8.91 mL/min/1.73 m².

Consistent with the 4C cohort, the iScore stratified risk of the composite kidney endpoint in a stepwise manner (Fig. 4A). Overall event rates were lower than in 4C, most likely reflecting the higher baseline kidney function and adult population-based design of the UKBB cohort. The iScore remained independently associated with the composite kidney endpoint in Cox proportional hazards models adjusted for available clinical covariates (Fig. 4B). UACR was not included in these models because it was available only for a limited subset of participants.

**Figure 4.**
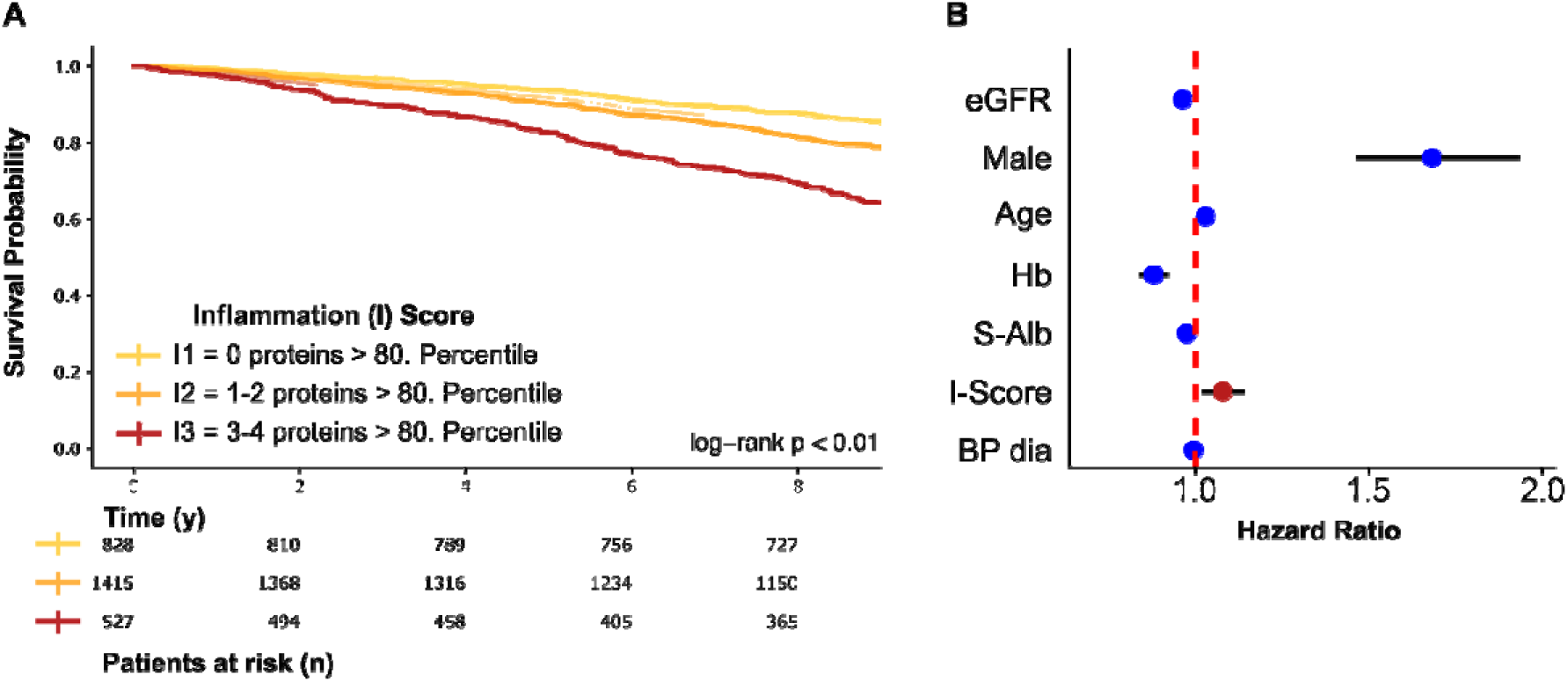
External evaluation of the inflammation-based risk score in the UK Biobank. (A) Kaplan–Meier analysis for the composite kidney endpoint according to inflammation score (iScore) category in UK Biobank participants with baseline eGFR <60 mL/min/1.73m². Numbers at risk are shown below the plot. P-value was calculated by log-rank test. (B) Multivariable Cox proportional hazards model for the composite kidney endpoint in the UK Biobank. Points indicate hazard ratios and horizontal lines indicate 95% confidence intervals.

### Single-cell analyses link serum inflammatory markers to intrarenal pathway activation in CKD

To place the serum-derived inflammatory signature into a kidney tissue context, we reanalysed a published single-cell transcriptomic atlas comprising 51 healthy and 53 CKD kidney samples, corresponding to approximately 92,000 and 178,000 single-cell transcriptomes, respectively^19^. We focused on the proteins included in the iScore together with their corresponding ligands and receptors. At the tissue level, transcripts encoding CD40, CD40LG, CD137/TNFRSF9, TNFSF9, and PD-1/PDCD1 were increased in CKD kidney tissue, whereas PD-L1/CD274 and CX3CL1 showed no overall increase (Fig. 5A). Cell compartment–resolved analysis revealed that kidney-infiltrating immune cells and epithelial cells showed the strongest upregulation of these axes, including PD-L1/CD274 and CX3CL1, while interstitial cells showed increased TNFRSF9 expression and endothelial cells showed lower expression of several of the corresponding genes in CKD (Fig. 5B).

**Figure 5.**
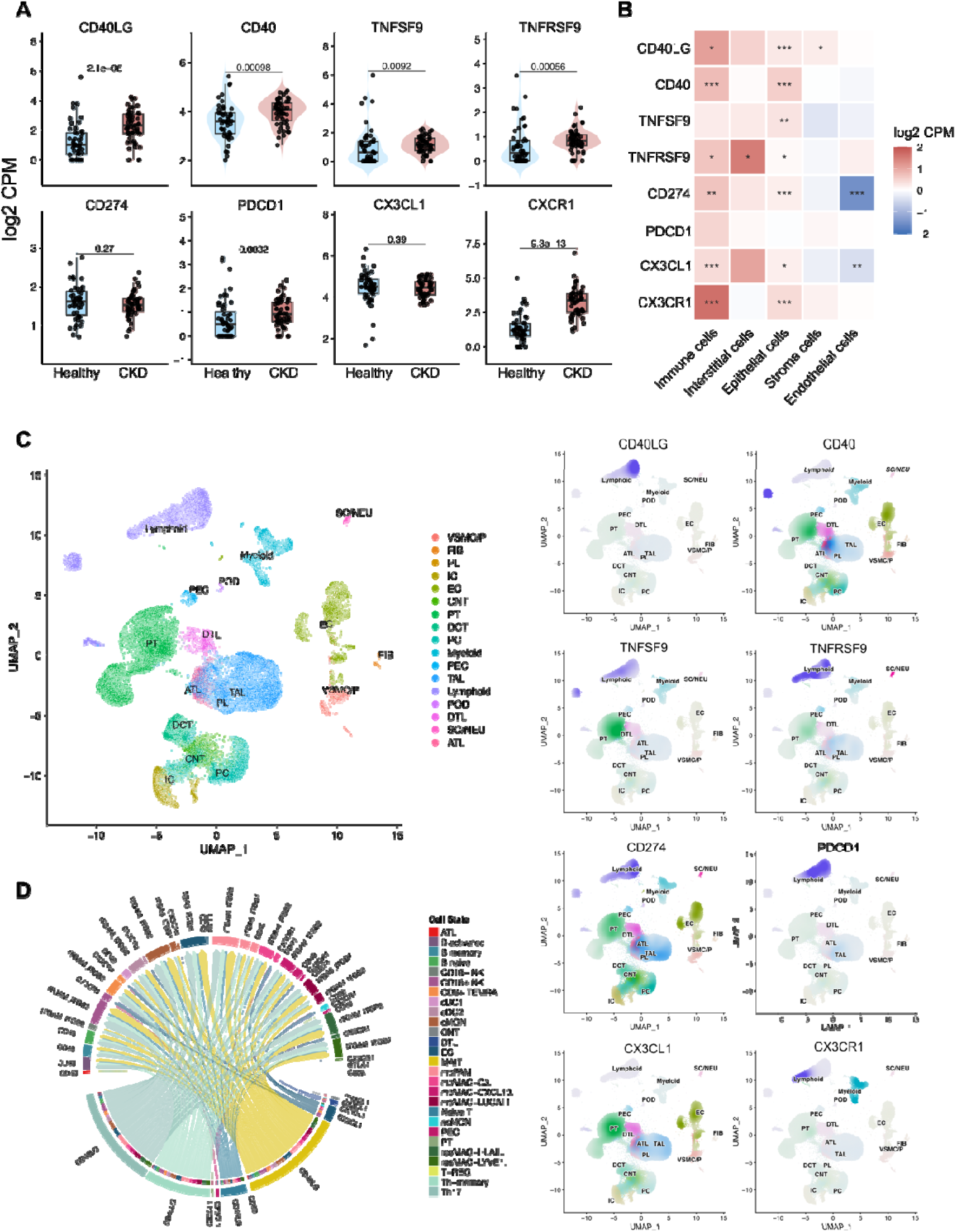
Single-cell analyses link serum inflammatory markers to intrarenal immune–parenchymal signaling in CKD. (A) Pseudobulk expression of genes encoding iScore-associated proteins and their corresponding ligand–receptor partners in healthy and CKD kidney tissue. Violin plots show sample-level distributions; P values were calculated by Wilcoxon rank-sum test. (B) Cell compartment–resolved differential expression of iScore-associated ligand–receptor axes in CKD compared with healthy kidneys. Color indicates log_2_-transformed fold change; asterisks indicate adjusted P values. (C) Uniform manifold approximation and projection (UMAP) embedding of CKD kidney single-cell transcriptomes annotated by renal and immune cell populations. (D) UMAP feature plots showing expression of CD40LG, CD40, TNFSF9, TNFRSF9, CD274, PDCD1, CX3CL1, and CX3CR1 across CKD kidney cell populations. (E) CellChat-based inference of ligand–receptor communication involving iScore-associated axes in CKD kidney tissue. Chord plot shows predicted interactions between immune and kidney-resident cell populations.

Mapping gene expression onto the CKD single-cell embedding indicated that ligands and receptors were distributed across distinct immune and non-immune cell populations rather than confined to a single cell lineage (Fig. 5C,D). We therefore inferred ligand–receptor signaling using CellChat to assess whether these axes may contribute to intrarenal cell–cell communication. This analysis recovered CD40LG–CD40, CX3CL1–CX3CR1, and PD-1/PD-L1-related signaling, with predicted interactions involving kidney-infiltrating immune cells as well as resident epithelial, endothelial, and interstitial compartments (Fig. 5E). Together, these analyses provide tissue-level support for the biological relevance of the serum-derived inflammatory signature and suggest that the iScore-associated proteins reflect immune–parenchymal communication within the injured kidney.

## Discussion

In this study, we provide a comprehensive analysis of the inflammation-related serum proteome in children with CKD and identify kidney function (eGFR, UACR) as dominant serum proteome determinant. Beyond descriptive profiling, we define a four-protein inflammation score (iScore) that refines prediction of CKD progression beyond eGFR and UACR^20^ in the 4C cohort and shows supportive external evidence in the UK Biobank.

By leveraging the well-characterized prospective 4C cohort^16^, we demonstrate that kidney function is the dominant determinant of variance in the circulating inflammatory proteome. This observation is supported by multiple analytical approaches, including PCA, multivariable linear modeling and confounder-aware association analyses. So far, inflammation in CKD has been described largely on single protein levels^6,21,22^, and a comprehensive analysis is still missing. The use of a pediatric cohort is a key strength of this study. In contrast to adult CKD populations, common CKD-related comorbidities such as diabetes, long-standing hypertension, and metabolic syndrome, which are known to independently modulate systemic inflammation, are usually absent^11,16^. This confounder-reduced setting enables a clear attribution of inflammatory signatures to kidney dysfunction itself. Our findings therefore provide important evidence that CKD is intrinsically linked to a systemic inflammatory state, rather than merely amplifying inflammation secondary to comorbidities. These findings are in line with observed inflammatory states in experimental CKD models such as subtotal nephrectomy, where the loss of kidney mass is intrinsically linked to SCI^8,9,23^.

In addition to the descriptive analysis of inflammation in pediatric CKD, we identified a subset of inflammatory proteins that are consistently associated with kidney outcomes over time, even after rigorous adjustment for established risk factors. Notably, proteins such as IL-15RA, VEGFA, PD-L1, FGF-23, CD40, CD137, and CX3CL1 emerged as key candidates with predictive value for CKD progression. Our data confirm reported markers for CKD progression from the pediatric CKiD cohort, like FGF-23^18^. We therefore focused on the newly identified markers CX3CL1, PD-L1, CD40, and CD137. Among the four proteins prioritized for the iScore, CX3CL1 points to chemokine-driven leukocyte recruitment and endothelial–immune crosstalk, whereas PD-L1, CD40, and CD137 implicate immune-regulatory and co-stimulatory signaling pathways. CX3CL1/CX3CR1 signaling has repeatedly been linked to renal inflammation and fibrosis, supporting its biological relevance in CKD^24,25^. CD40/CD40L signaling is likewise implicated in renal injury and has attracted interest as a therapeutic target in kidney disease^26,27^. CD137 has been linked to macrophage activation and kidney fibrosis in experimental CKD^28^. PD-L1 is more difficult to interpret in a kidney-specific context, but may reflect broader immune-regulatory states associated with chronic tissue injury and senescence^29^. Together, these observations support the interpretation that the iScore reflects biologically meaningful inflammatory signaling rather than nonspecific retention of circulating proteins.

We aimed to derive a score that captures inflammatory activity beyond the dominant effects of eGFR and UACR. To this end, elevated protein levels were defined from residualized values adjusted for kidney function parameters, conceptually analogous to age-adapted percentile approaches used in pediatrics and to biomarker interpretation frameworks that account for kidney function^30^. This strategy allowed us to compare patients with broadly similar conventional CKD risk profiles but clearly different outcomes, underscoring that the iScore captures prognostic information not contained in eGFR and UACR alone. Although this approach may be less optimized for internal prediction than purely data-driven model construction, it improves biological interpretability and may facilitate future clinical translation. Importantly, the relevance of the iScore was validated in the UK Biobank, where it stratified outcome despite major differences in age, cohort composition, and available covariates. Therefore, these analyses should be viewed as supportive external evaluation rather than definitive clinical validation^31^. Nevertheless, in line with a rare-to-common translational framework^32^, our data suggest that the inflammatory processes captured by the score may not be restricted to pediatric CKD, but could reflect more general pathways of age-overarching progressive kidney disease^6^.

An important question is whether these proteins primarily reflect reduced kidney clearance or active disease biology. By reanalysing kidney single-cell transcriptomic data, we mapped the iScore-associated ligand–receptor axes to intrarenal cell states. This analysis showed that CD40/CD40L, CD137/CD137L, CX3CL1/CX3CR1, and PD-1/PD-L1 signaling were represented across immune and non-immune compartments of the CKD kidney, supporting a role in immune–parenchymal crosstalk. In particular, the epithelial compartment emerged as a relevant non-immune site for several of these axes, whereas interstitial cells showed a more restricted signal, mainly involving CD137. CD40/CD40LG signaling in renal epithelial cells has been linked to NF-κB–dependent cytokine and chemokine production, and tubular CD40 expression correlates with inflammatory mediators and macrophage infiltration^33^. CD137/CD137L signaling provides a second example of immune–epithelial crosstalk, as CD137L signaling in tubular epithelial cells can induce CXCL1/CXCL2 production and neutrophil recruitment in experimental kidney injury^34^. Macrophage-derived CD137L has been implicated in renal lymphangiogenesis and fibrosis^28^. CX3CL1/CX3CR1 signaling further supports an immune–parenchymal interface by promoting recruitment and retention of CX3CR1-positive myeloid cells in kidney inflammation and fibrosis^35^. PD-L1 requires a more cautious interpretation: in tubular epithelial cells, PD-L1 can suppress T-cell cytokine production and may act as a counter-regulatory pathway limiting immune-mediated epithelial injury^36^, although persistent PD-L1 expression may also mark chronic injury or senescence-associated inflammatory states^29^. Together, these findings support the concept that the iScore captures systemic correlates of immune–parenchymal inflammatory programs in CKD rather than nonspecific retention of circulating proteins alone.

Clinically, the added value of the iScore may lie in refining risk stratification within existing eGFR- and UACR -based categories^20^ rather than replacing them. In this study, children with comparable baseline kidney function and UACR showed markedly different outcome trajectories according to inflammatory burden. Such a biomarker strategy could ultimately support closer monitoring, trial enrichment, or prioritization of patients for anti-inflammatory interventions once suitable therapies become available^37^. However, prospective validation, assay standardization, and demonstration of clinical utility will be required before implementation in routine care.

Immunosuppressants and immune-targeting biologicals are available, with anti-IL-6 currently being investigated in CKD^38-40^ to attenuate cardiovascular disease progression. However, doubts about their suitability as long-term treatments remain, as the targeted mechanisms are often broad and non-specific, and side effects (e.g. infections) may outweigh the benefits.

We will provide de-identified OLINK protein measurements together with selected clinical metadata to support reproducibility and reuse, including age, sex, eGFR, UACR, and underlying disease category, within the limits of patient data protection. This resource will enable independent exploration of individual proteins while protecting privacy in this pediatric cohort.

Nevertheless, several methodological considerations need to be discussed. First, the use of the OLINK proximity extension assay provides high sensitivity for low-abundance proteins that are difficult to quantify with conventional immunoassays^41^. However, the resulting protein expression represents relative quantification^42^.First, the use of the OLINK proximity extension assay provides high sensitivity for low-abundance proteins that are difficult to quantify with conventional immunoassays^41^. However, the resulting protein expression represents relative quantification^42^. Consequently, clinically actionable cut-offs cannot be directly derived from these data. Future studies incorporating calibrated assays with standard curves will be necessary to establish reference values and define thresholds for clinical decision-making. In addition, the iScore was intentionally restricted to four interpretable markers and was not designed as an exhaustive proteomic prediction model. Other proteins, including IL-15RA, VEGFA, and M-CSF, also showed prognostic associations and may contribute to distinct CKD endotypes. Future studies should test whether broader or disease-specific panels improve prediction, while maintaining interpretability. Moreover, while circulating protein levels may reflect systemic inflammation, they do not necessarily indicate causal involvement in disease progression. The observation that several of the identified proteins are linked to immune checkpoint regulation further raises intriguing mechanistic questions, as these pathways encompass both inhibitory (e.g., PD-1/PD-L1 axis) and stimulatory (e.g., CD40, CD137) signals that could differentially influence renal inflammation and fibrosis.

In conclusion, our study provides comprehensive evidence that CKD profoundly shapes the inflammatory serum proteome and identifies a set of inflammation-related proteins with independent prognostic relevance. The iScore integrates this information into an interpretable framework that refines risk prediction beyond established markers. While further work is needed to establish mechanistic links and enable clinical implementation, these findings highlight the potential of inflammation-based biomarkers to improve our understanding and management of CKD across the lifespan.

## Acknowledgement

We thank all patients who participated in this study.

4C Study Consortium: The principal investigators of the 4C Study included the following: Pediatric Nephrology, University Children’s Hospital Vienna, Vienna, Austria: Klaus Arbeiter, MD; Department of Pediatrics I, Medical University, Innsbruck, Austria: Alejandra Rosales, MD; University Hospital Motol, Prague, Czech Republic: Jiri Dusek, MD; Pole Medico-Chirugical de Pediatrie, Hôpital de Hautepierre, Strasbourg, France: Ariane Zaloszyc, MD; Pediatric Nephrology, Charité Campus Virchow-Klinikum, Berlin, Germany: Uwe Querfeld, MD, and Jutta Gellermann, MD; Pediat-ric Nephrology Immunology and Hypertensiology, University Children’s Hospital, Cologne, Germany: Max Liebau, MD, and Lutz Weber, MD; Pediatric Nephrology, University Children’s Hospital, Erlan-gen, Germany: Evelin Muschiol, MD; Pediatric Nephrology, University Children’s Hospital, Essen, Germany: Rainer Büscher, MD; Pediatric Nephrology, UKE University Children’s Hospital, Hamburg, Germany: Jun Oh, MD; Pediatric Nephrology, Hannover Medical School, Hannover, Germany: Anette Melk, MD, Daniela Thurn-Valassina, MD, and Dieter Haffner, MD; Division of Pediatric Nephrol-ogy, Center for Children and Adolescents, Heidelberg, Germany: Franz Schaefer, MD; Division of Pediatric Nephrology, Center for Pediatrics and Adolescent Medicine, Freiburg, Germany: Charlotte Gimpel, MD; Division of Pediatric and Nephrology, Center for Pediatrics and Adolescent, Jena, Germany: Ulrike John, MD; Children’s Dialysis Center, Town Hospital St Georg, Leipzig, Germany: Simone Wygoda MD; KfH Kidney Center for Children, Marburg, Germany: Nikola Jeck, MD; Pediatric Nephrology, University Children’s Hospital, Rostock, Germany: Marianne Wigger, MD; Pediatric Nephrology and Dialysis, Ospedale Maggiore, Policlinico, Milano, Italy: Sara Testa, MD; Dialysis and Transplantation, Department of Pediatrics, Dipartimento di Pediatria Salus Pueri, Padova, Italy: Luisa Murer, MD; Division di Nefrologia e Dialisi, Ospedale Pediatrico Bambino Gesú, Roma, Italy: Chiara Matteucci, MD; Center for Pediatrics, University Children’s Hospital, Vilnius, Lithuania: Augustina Jankauskiene, MD, and Karolis Azukaitis, MD; Dialysis Unit, PA Children’s Hospital, Cracow, Poland: Dorota Drozdz, MD; Pediatric Nephrology, Instituto Giannina Gaslini, Genova, Italy: Francesca Lugani, MD; Department of Pediatric and Adolescent Nephrology, University Medical School, Gdańsk, Poland: Aleksandra Zurowska, MD; Pomeranian Academy of Medicine, Clinic of Pediatrics, Szczecin, Poland: Marcin Zaniew, MD; Nephrology, Kidney, Transplantation and Hypertension, Children’s Memorial Health Institute, Warsaw, Poland: Mieczyslaw Litwin, MD, and Anna Nimierska, MD; Serviçio de Pediatria, Hospital de São João, Porto, Portugal: Ana Teixeira, MD; University Children’s Hospital, Belgrade, Serbia: Amira Peco-Antic, MD, and Dusan Paripovic, MD; Nephrology, University Children’s Hospital, Zürich, Switzerland: Guido Laube, MD; Cocuk Nefrolojisi Bilim Dali, Cukurova Universitesi Tip Fakültesi, Adana, Turkey: Ali Anarat, MD, and Aysun Bayazit, MD; Pediatric Nephrology, Hacettepe University, Ankara, Turkey: Ali Duzova, MD, and Yelda Bilginer, MD; Pediat-ric Nephrology, Cerrahpasa Medical Faculty, Istanbul, Turkey: Salim Caliskan, MD, Nur Canpolat, MD, and Mahmut Civilibal, MD; Pediatric Nephrology, Ege University, Izmir, Turkey: Sevgi Mir, MD, and Betül Sözeri, MD; Pediatric Nephrology, University Children’s Hospital, Münster, Germany: Brigitta Kranz, MD; Department of Pediatrics, Sant’Orsola-Malphighi Hospital, Bologna, Italy: Francesca Mencarelli, MD; Inselspital Children’s Hospital, Bern, Switzerland: Brigitte Dorn, MD; Pediatric Nephrology, University Tip Faculty of Medicine Cebeci Kampusu Cocuk, Ankara, Turkey: Fatos Yalcinkaya, MD; Faculty of Medicine, Baskent University, Ankara, Turkey: Esra Baskin, MD; Diskapi Children’s Hospital, Ankara, Turkey: Nilgun Cakar, MD; Pediatric Nephrology, Gazi University Hospital, Ankara, Turkey: Oguz Soylemezoglu, MD; Department of Pediatric Nephrology, Istanbul Medical Faculty, Çapa-Istanbul, Turkey: Sevinc Emre, MD; Pediatric Nephrology, Goztepe Educational and Research Hospital, Istanbul, Turkey: Cengiz Candan, MD; Pediatric Nephrology, Bakirkoy Children’s Hospital, Istanbul, Turkey: Aysel Kiyak, MD; Pediatric Nephrology, Sisli Educational and Research Hospital, Istanbul, Turkey: Gul Ozcelik, MD; Pediatric Nephrology, Marmara University Medical Faculty, Istanbul, Turkey: Harika Alpay, MD; Nephrology, Great Ormond Street Hospital, London, United King-dom: Rukshana Shroff, MD; Service de Néphrologie Pédiatrique, Universite Hôpital Femme Mère Enfant, Lyon, France: Bruno Rachin, MD; Service de Pédiatrie, Centre de Reference Maladies Re-nales Rares du Sud-Quest, Bordeaux, France: Jerome Harambat, MD; Pediatric Nephrology, Zabrze, Poland: Maria Szczepanska, MD; Dortcelik Children’s Hospital, Bursa, Turkey: Hakan Erdogan, MD; Uludag University, Bursa, Turkey: Osman Donmez, MD; Department of Pediatric Nephrology, University of Gaziantep, Gaziantep, Turkey: Ayse Balat, MD; Tepecik Training and Research Hospital, Izmir, Turkey: Nejat Aksu, MD; Department of Pediatric Nephrology, Inonu Universtiy Medical School, Malatya, Turkey : Yilmaz Tabel, MD; Pediatric Nephrology Department, Celal Bayar University, Manisa, Turkey: Pelin Ertan, MD; and Pediatric Nephrology, Sanliurfa Children’s Hospital, Sanliurfa, Tur-key: Ebru Yilmaz, MD.

## Data availability statement

We provide all our proteomics data as an open-access app for exploration: https://hbartolomaeus.shinyapps.io/olink_inflammation_4c/. Deidentified proteomics data and metadata, including age, sex, eGFR and UACR, can be obtained from Zenodo (10.5281/zenodo.20622389). Identification and additional metadata can be obtained from the corresponding authors after obtaining consent from the 4C steering committee.

