## Supplement for "Inflammation-based risk score predicts kidney survival in children and adults with chronic kidney disease"

**Supplemental Methods**

All data processing and analysis were done with R (version 4.5.0). The R-scripts are available on GitHub (<https://github.com/rosareitmeir/4C_OLINK_CKDprogression>).

**Linear Models and Lasso Regression**

For linear models, we used the R *stats* package (version 4.5.0). Predictor significance in linear models was determined using two-sided t-tests on the regression coefficients. In cases with multiple coefficients, the resulting p-values were corrected accordingly, using the Benjamini-Hochberg procedure.

LASSO regression was implemented with the *glmnet* package (version 4.1-10). Missing values are imputed with the median. The different variables are grouped in five different meta categories: 1: kidney function with the variables of eGFR, UACR and CKD stage, 2: blood pressure: numeric variables of systolic and diastolic sds values and their binary counterpart whether the sds value is above 2 and hypertension group, 3: underlying disease, 4: country of origin, and 5: demographics including age, sex, and BMI sds.

For each combination of protein and variable category, we first identified the optimal regularization strength (lambda) by performing 10-fold-cross validation using LASSO regression, minimizing mean squared error. Using the selected lambda, we then fitted the final LASSO model. Model fit was evaluated based on the adjusted R² metric. P-values were corrected for multiple testing using the Benjamini-Hochberg procedure, with a false discovery rate (FDR) threshold of 0.05.

**Contrasts**

To assess the impact of diagnosis and country of origin, Type 2 ANCOVA was performed to test for differences between the subgroups while adjusting for eGFR. ANCOVA was conducted using the *rstatix* package (version 0.7.3). For proteins with a significant ANCOVA result (FDR adjusted p <0.05), post-hoc contrast analyses were performed using the *emmeans* package (version 2.0.0). For this, linear models were built, including diagnose or country, along with eGFR, age, sex, and BMI sds as covariates. The p-values of the contrasts are corrected using the Tukey method, with adjusted p-value <0.05 considered significant. Standardized effect sizes were reported for each contrast.

**Confounding-Aware Statistical Testing**

For univariate statistical tests, *metadeconfoundR* (version 1.0.2) was used, which performs Wilcoxon rank sum test for binary variables and Spearman's correlation for continuous variables. Additionally, the tool tests for any (univariate) confounding. In total 18 possible covariates were considered (see **Clinical Variable List**). P-values were adjusted by the Benjamini-Hochberg procedure. An association was considered significant when the adjusted p-value was <0.05. Effect sizes were reported as Cliff’s Delta.

**Random Forest**

Random Forests were built using the package *randomForest* (version 4.7-1.2). Features for the Random Forests were KFRE variables (eGFR, UACR, age, sex), all 77 proteins levels, and clinical variables including BMI sds, underlying disease, blood values, blood pressure and hypertension related variables (see **Clinical Variable List**), are functioning as features for the Random Forests. Missing values were imputed with the median. The data set was split 80 to 20 into train and test data. To further prevent overfitting, a k-cross validation was implemented using a fold size of 5. The random forests were evaluated on the test data, calculating sensitivity, specificity, F-measure, and ROC-curves. Feature importance was assessed by mean decrease accuracy.

**Survival Analysis**

Cox-Proportional Hazard Models were built with the *survival* package (version 3.8-3), and Kaplan-Meier Curves were calculated by the *survminer* package (version 0.5.1). We defined the survival time as the time passed from the first visit till reaching the composed endpoint in years, using the recorded age of the patients at the time point of the transplantation, start of dialysis or death. For patients having a 50% eGFR loss, linear interpolation is used to estimate the age point when the 50% loss is reached. For this, all available eGFR measurements were used between the first visit and the visit when the loss was detected. Patients with no follow-up visits and not reaching the composite endpoint were excluded from this analysis.

The assumption of proportional hazard was visually inspected by plotting the Schonefeld residuals and formally tested using the test of Grambsch and Therneau^1^ implemented in the *cox.zph* function. As eGFR and age were not proportional over time, the models are stratified by the CKD stages (3a,3b,4,5) and in two age groups, split at the median age of 12.2 years at study entry. CKD stage 2 is removed from this analysis, as the number of patients was not sufficient for its own strata (n=8).

To investigate the impact of systemic inflammation on survival, we defined an inflammation score as follows. For each of the four proteins of interest, we fitted a linear model with protein level as the outcome and eGFR and UACR as predictors. We extracted the residuals from these models, which represent the variation in protein levels not explained by kidney function. Then, for every sample, we counted the number of residuals exceeding the 80^th^ percentile. This sum, ranging from zero to four, was defined as the inflammation score. The inflammation score was used as a continuous predictor in the Cox regression and categorized into three groups (0,1/2,3/4 high residuals) for Kaplan-Meier survival curves.

The significance of predictors in the Cox model was assessed using Wald tests. Results are reported as hazard ratios (exponentiated regression coefficients) along with their 95% confidence intervals. Kaplan-Meier survival curves were compared using the log-rank test.

**Slope Prediction**

To estimate eGFR slope, we calculated the difference between eGFR at study entry and at the 12-month visit. Patients without an eGFR value at month 12 were excluded from the analysis. For patients receiving RRT at 12 months, the 12-month eGFR value was imputed with ten. To account for varying baseline CKD stages, the relative eGFR change was computed by dividing the absolute difference by the baseline eGFR.

**Protein-Protein Partial Correlation Network Analysis**

We computed partial correlations for all protein-protein pairs using the package *ppcor* (version 1.1), while controlling for all other proteins. To additionally account for kidney function, partial correlations were calculated on the residuals of linear models with eGFR and UACR as explanatory variables. The resulting network was visualized using the package *ggraph* (version 2.2.2), representing proteins as nodes and significant partial correlations (FDR adjusted p < 0.05) as edges.

**Clinical Variable List**

- Age in years at study entry
- Sex
- eGFR at study entry: Zappitelli formula: given creatinine (crea), cystatin C (cystc) and blood urea nitrogen (bun), eGFR is calculated as follows 39.1*((height/100)/creat)**0.516 * (1.8/cystc)**0.294 * (30/bun)**0.169 * ((height/100)/1.4)**0.188, for male patients eGFR is multiplied by 1*.099.
- BMI sds at study entry
- Underlying disease categorized in CAKUT, Glomerulopathies, post AKI/HUS, Tubint., and other disease
  - CAKUT: Renal Hypo-/Dysplasia, Reflux nephropathy, Obstructive nephropathy, Syndromal malformation of urinary tract, ADPKD, ARPKD, Dystopia, Malposition, Aplasia/Agenesis, Hypoplasia, Renal dysplasia, Multicystic dysplasia, Segmental dysplasia, Oligomeganephronia, Ureter of upper renal pelvis inserting caudally, Stenosis of pyelo-ureteral junction, Vesicoureteral reflux, Ureter fissus of duplex, Megaureter, Ectopic ureter, Ureterocele, Bladder anomalies, Urethral anomalies, Other renal or urinary tract malformation
  - Glomerulopathies: IgA nephropathy, ANCA-associated glomerulonephritis, IgA vasculitis with glomerulonephritis, Systemic lupus erythematosus, Congenital nephrotic syndrome, Infantile nephrotic syndrome, Syndromal nephrotic syndrome, Minimal-change glomerulopathy, Focal segmental glomerulosclerosis, Membranous glomerulopathy, Mesangioproliferative nephropathy, Membranoproliferative glomerulonephritis, Rapidly progressive glomerulonephritis, Post-infectous glomerulonephritis, Alport Syndrome
  - Post AKI/HUS: Post-ischemic chronic renal failure, Hemolytic uremic syndrome
  - Tubint.: Interstitial nephropathy, Cystinosis, Oxalosis, Nephrocalcinosis, Nephronophthisis, Metabolic
- UACR, log-transformed, ratio of urinary albumin to urinary creatinine times 100, at study entry
- Serum albumin in g/L at study entry
- Bicarbonate in mmol/L at study entry (blood)
- Hemoglobin in g/dL at study entry (blood)
- Phosphorus in mmol/L at study entry (blood)
- Office diastolic and systolic sds values, and dia./sys. sds >2 as binary variables,
- Hypertension, binary, defined as either having antihypertensive treatment or increased blood pressure measurements at study entry
- Refined Hypertension category:
  - Antihypertensive treatment and controlled blood pressure
  - No antihypertensive treatment and normal blood pressure
  - Antihypertensive treatment and uncontrolled blood pressure
  - No antihypertensive treatment and uncontrolled blood pressure
- Intake of RAS inhibitors at study entry
- Intake of Immunosuppressiva at study entry
- Country of origin: Austria, France, Germany, Italy, Poland, Serbia, Turkey, UK, or other country
- Type of CKD endpoint: Transplantation, Dialysis, or 50% eGFR loss since study entry

**Analysis of the UK Biobank Data**

Study participants

The UK Biobank is a population-based cohort of 502,366 participants aged 40–69 years recruited between 2006 and 2010 across 22 assessment centres in England, Scotland, and Wales. Ethical approval was granted by the North West Multi-Centre Research Ethics Committee (reference 11/NW/0382), and all participants provided written informed consent. Data available as of 16 November 2023 were used for this study.

Dialysis, kidney transplantation, and CKD stages were identified using International Classification of Diseases, Tenth Revision (ICD-10) diagnosis codes and the Observational Medical Outcomes Partnership (OMOP) Common Data Model. For each participant, the earliest recorded occurrence of each event was captured. Dialysis event was determined by consolidating the ICD-10 codes Z99.2, Z49.0-Z49.2, T82.4, and Y84.1. Kidney transplantation was defined using the codes Z94.0 and T86.1, while CKD stages 1-5 were captured using N18.1-N18.5.

Clinical data, proteomics analysis and inflammation score

eGFR was derived from plasma cystatin C concentrations using the equation eGFR = 74.835 / (cystatin C¹·³³³). OLINK targeted proteomics was measured as described previously^2^. Plasma levels of four inflammation-related proteins—CD40, CD137 (TNFRSF9), CX3CL1, and PD-L1 (CD274)—were extracted and examined for their association with kidney outcomes. To minimize confounding by baseline kidney function, protein concentrations were residualized against eGFR by fitting linear regression models for each protein. For each participant, proteins with residuals exceeding the 80^th^ percentile were classified as elevated. Based on the number of elevated proteins, participants were assigned an inflammation score: I1, 0 proteins above the 80^th^ percentile; I2, 1–2 proteins above the 80^th^ percentile; and I3, 3–4 proteins above the 80^th^ percentile.

eGFR decline assessment

As individual follow-up eGFR measurements were unavailable, kidney function decline was inferred from diagnostic codes corresponding to CKD stages based on ICD and OMOP mappings. Each CKD stage represents an eGFR interval (e.g., Stage 4 = 15–29 mL/min/1.73 m²), introducing uncertainty regarding the precise eGFR value. Relative eGFR decline was defined as the difference between baseline eGFR (${eGFR}_{BL}$) and the upper limit of the eGFR range corresponding to the first observed follow-up CKD stage (${eGFR}_{FU}^{UL}$), normalized to baseline eGFR. This definition yields a conservative (upper-bound) estimate of eGFR decline, as the true follow-up eGFR may be lower.

$$Relative eGFR decline = \frac{{eGFR}_{BL} - {eGFR}_{FU}^{UL}}{{eGFR}_{BL}}$$

The composite kidney endpoint was defined as the occurrence of at least one of the following events: 50% eGFR decline, dialysis, kidney transplantation, or death. For participants experiencing multiple qualifying events, the earliest recorded date was used to assign the time of outcome occurrence.

Statistical analysis

Kidney survival was assessed using Kaplan–Meier curves, with time from baseline to the first occurrence of each composite outcome. Differences across inflammation groups were compared using the log-rank test. Kaplan-Meier analysis was restricted to participants with eGFR < 60 mL/min/1.73 m² at baseline.

Continuous variables were evaluated for normality using the Shapiro–Wilk test (α > 0.05). If all groups met the normality assumption, mean differences were tested using linear regression (ANOVA). Otherwise, the non-parametric Mann–Whitney U-test was applied. For categorical variables, group differences were assessed using Pearson’s χ² test or Fisher’s exact test, as appropriate. Benjamini–Hochberg FDR method adjusted for multiple testing. Analyses were conducted in Python using statsmodels, scipy, lifelines and pandas.

**Analysis of the kidney single cell data**

We analysed human kidney single-cell RNA-seq data from the Kidney Precision Medicine Project^3^. The processed Seurat object was imported into R (v4.5.1) using Seurat (v5.4.0), no reannotation or re-integration was performed. For the iScores ligand-receptor pairs, pseudobulk profiles were generated by summing raw counts per specimen, normalizing to counts per million and log-transforming. Expression was compared between CKD and healthy specimens for each gene using two-sided Wilcoxon rank-sum tests, shown as violin plots, and the same comparison was repeated within each cell class and displayed as a gene-by-class heatmap. Within CKD cells, per-transcript abundance across the UMAP was estimated by weighted kernel density estimation (ks) to color cells by expression intensity. Ligand-receptor signaling was inferred with CellChat and visualized as chord diagrams. Visualizations used ggplot2 (v4.0.2) and ggpubr (v0.6.3).

**Supplemental Tables**

**Table S1.** Results of random forest validation in test data for different cross-sectional endpoints (month).

| **RF_model** | **Sensitivity** | **Specificity** | **Accuracy** | **Precision** | **F_Measure** | **Month** |
| --- | --- | --- | --- | --- | --- | --- |
| **KFRE** | 0,429 | 0,954 | 0,868 | 0,643 | 0,514 | 12 |
| **All** | 0,048 | 1,000 | 0,845 | 1,000 | 0,091 | 12 |
| **Proteome** | 0,048 | 1,000 | 0,845 | 1,000 | 0,091 | 12 |
| **KFRE** | 0,632 | 0,907 | 0,814 | 0,774 | 0,696 | 24 |
| **All** | 0,579 | 0,920 | 0,805 | 0,786 | 0,667 | 24 |
| **Proteome** | 0,421 | 0,987 | 0,796 | 0,941 | 0,582 | 24 |
| **KFRE** | 0,698 | 0,814 | 0,765 | 0,732 | 0,714 | 36 |
| **All** | 0,674 | 0,847 | 0,775 | 0,763 | 0,716 | 36 |
| **Proteome** | 0,558 | 0,847 | 0,725 | 0,727 | 0,632 | 36 |
| **KFRE** | 0,830 | 0,596 | 0,713 | 0,672 | 0,743 | 48 |
| **All** | 0,872 | 0,617 | 0,745 | 0,695 | 0,774 | 48 |
| **Proteome** | 0,681 | 0,660 | 0,670 | 0,667 | 0,674 | 48 |
| **KFRE** | 0,800 | 0,545 | 0,736 | 0,839 | 0,819 | 60 |
| **All** | 0,877 | 0,591 | 0,805 | 0,864 | 0,870 | 60 |
| **Proteome** | 0,846 | 0,364 | 0,724 | 0,797 | 0,821 | 60 |
| **KFRE** | 0,906 | 0,368 | 0,783 | 0,829 | 0,866 | 72 |
| **All** | 0,938 | 0,316 | 0,795 | 0,822 | 0,876 | 72 |
| **Proteome** | 0,969 | 0,158 | 0,783 | 0,795 | 0,873 | 72 |
| **KFRE** | 0,939 | 0,308 | 0,835 | 0,873 | 0,905 | 84 |
| **All** | 1,000 | 0,000 | 0,835 | 0,835 | 0,910 | 84 |
| **Proteome** | 0,985 | 0,000 | 0,823 | 0,833 | 0,903 | 84 |
| **KFRE** | 0,985 | 0,111 | 0,878 | 0,889 | 0,934 | 96 |
| **All** | 1,000 | 0,000 | 0,878 | 0,878 | 0,935 | 96 |
| **Proteome** | 0,985 | 0,000 | 0,865 | 0,877 | 0,928 | 96 |

**Table S2.** Baseline characteristics of the UK Biobank participants with available CD40, CD137, CX3CL1, PD-L1, and eGFR measurements. Values are shown as mean (standard deviation) for continuous variables and n (%) for categorical variables. P-values were calculated using linear regression for continuous and χ² tests for categorical variables.

|  | **eGFR <60 (n=2,770)** | **eGFR >60 (n=44,952)** | **p-value** |
| --- | --- | --- | --- |
| Age at recruitment (years) | 62.32 (6.19) | 56.44 (8.21) | <0.001 |
| Male sex | 1,595 (57.6%) | 20,369 (45.3%) | <0.001 |
| BMI (kg/m²) | 30.64 (6.04) | 27.26 (4.63) | <0.001 |
| eGFR (mL/min/1.73 m²) | 51.18 (8.91) | 90.67 (16.91) | <0.001 |
| Waist circumference (cm) | 100.62 (14.82) | 89.78 (13.10) | <0.001 |
| Mortality (all-cause) | 1,010 (36.5%) | 4,141 (9.2%) | <0.001 |
| HFpEF | 739 (26.7%) | 2,251 (5.0%) | <0.001 |
| Pulse rate (bpm) | 71.55 (13.42) | 69.38 (11.20) | <0.001 |
| Systolic blood pressure (mmHg) | 141.58 (19.77) | 137.50 (18.53) | <0.001 |
| Diastolic blood pressure (mmHg) | 81.89 (11.21) | 82.17 (10.12) | 0.168 |
| Pulse pressure (mmHg) | 59.69 (15.71) | 55.34 (13.50) | <0.001 |
| Sleep apnoea | 39 (1.4%) | 196 (0.4%) | <0.001 |
| Chronic ischaemic heart disease | 393 (14.2%) | 1,333 (3.0%) | <0.001 |
| Nonrheumatic mitral valve disease | 45 (1.6%) | 124 (0.3%) | <0.001 |
| Nonrheumatic aortic valve disease | 28 (1.0%) | 87 (0.2%) | <0.001 |
| Cardiomyopathy | 35 (1.3%) | 72 (0.2%) | <0.001 |
| Varicose veins of lower extremities | 58 (2.1%) | 817 (1.8%) | 0.332 |
| Hypotension | 32 (1.2%) | 126 (0.3%) | <0.001 |
| Angina pectoris/ coronary artery disease | 280 (10.1%) | 1,058 (2.4%) | <0.001 |
| Endocrine, nutritional and metabolic diseases | 763 (27.5%) | 3,249 (7.2%) | <0.001 |
| Mental and behavioral disorders | 189 (6.8%) | 1,032 (2.3%) | <0.001 |
| Diseases of the nervous system | 319 (11.5%) | 2,400 (5.3%) | <0.001 |
| Diseases of the eye and adnexa | 277 (10.0%) | 2,019 (4.5%) | <0.001 |
| Diseases of the respiratory system | 507 (18.3%) | 3,292 (7.3%) | <0.001 |
| Diseases of the digestive system | 1,094 (39.5%) | 9,639 (21.4%) | <0.001 |
| Diseases of the skin and subcutaneous tissue | 322 (11.6%) | 2,557 (5.7%) | <0.001 |
| Diseases of the musculoskeletal system and connective tissue | 883 (31.9%) | 6,536 (14.5%) | <0.001 |
| Diseases of the genitourinary system | 772 (27.9%) | 7,108 (15.8%) | <0.001 |
| Pregnancy, childlbirth and the puerperium | 10 (0.4%) | 1,693 (3.8%) | <0.001 |
| Congenital malformations, deformations and chromosomal abnormalities | 54 (1.9%) | 311 (0.7%) | <0.001 |
| Aortic aneurysm | 26 (0.9%) | 42 (0.1%) | <0.001 |
| Phlebitis and thrombophlebitis | 83 (3.0%) | 501 (1.1%) | <0.001 |
| Type 2 diabetes | 377 (13.6%) | 1,260 (2.8%) | <0.001 |
| Myocardial infarction | 188 (6.8%) | 680 (1.5%) | <0.001 |
| Chronic kidney disease | 461 (16.6%) | 495 (1.1%) | <0.001 |
| Hypertensive renal disease | 121 (4.4%) | 22 (0.0%) | <0.001 |
| Endocarditis | 19 (0.7%) | 132 (0.3%) | <0.001 |
| Essential (primary) hypertension | 929 (33.5%) | 3,354 (7.5%) | <0.001 |
| Atrial fibrillation and flutter | 218 (7.9%) | 847 (1.9%) | <0.001 |
| Cerebrovascular event/ stroke | 133 (4.8%) | 466 (1.0%) | <0.001 |
| Embolism and thrombosis | 156 (5.6%) | 925 (2.1%) | <0.001 |
| Pulmonary embolism | 57 (2.1%) | 209 (0.5%) | <0.001 |
| Pulmonary artery hypertension | 42 (1.5%) | 117 (0.3%) | <0.001 |
| Neoplasms | 612 (22.1%) | 6,692 (14.9%) | <0.001 |
| Congenital malformations of the circulatory system | 16 (0.6%) | 92 (0.2%) | <0.001 |
| Seizure/epilepsy | 46 (1.7%) | 519 (1.2%) | 0.023 |
| Swollen ankle region | 72 (2.6%) | 408 (0.9%) | <0.001 |
| Impaired exercise tolerance | 114 (4.1%) | 823 (1.8%) | <0.001 |
| Tachycardia | 12 (0.4%) | 49 (0.1%) | <0.001 |
| Bradycardia | 19 (0.7%) | 96 (0.2%) | <0.001 |
| Palpitations | 32 (1.2%) | 206 (0.5%) | <0.001 |
| Cough | 27 (1.0%) | 135 (0.3%) | <0.001 |
| Symptoms and signs involving the digestive system and abdomen | 378 (13.6%) | 3,345 (7.4%) | <0.001 |
| Symptoms and signs involving the skin and subcutaneous tissue | 55 (2.0%) | 399 (0.9%) | <0.001 |
| Symptoms and signs involving the urinary system | 238 (8.6%) | 1,592 (3.5%) | <0.001 |
| Symptoms and signs involving cognition, perception, emotional state and behaviour | 58 (2.1%) | 329 (0.7%) | <0.001 |
| Symptoms and signs involving speech and voice | 23 (0.8%) | 154 (0.3%) | <0.001 |
| Dyspnoea | 262 (9.5%) | 1,540 (3.4%) | <0.001 |
| General, local, unspecified oedema | 38 (1.4%) | 41 (0.1%) | <0.001 |
| Nocturia | 22 (0.8%) | 216 (0.5%) | 0.034 |
| Post-viral fatigue | 12 (0.4%) | 206 (0.5%) | 0.964 |
| Syncope | 108 (3.9%) | 880 (2.0%) | <0.001 |
| Fatigue (excl. post-viral) | 173 (6.2%) | 2,059 (4.6%) | <0.001 |
| Cholesterol lowering medication | 1,237 (44.7%) | 7,647 (17.0%) | <0.001 |
| Mineralocorticoid receptor antagonist | 55 (2.0%) | 61 (0.1%) | <0.001 |
| Levothyroxine | 105 (3.8%) | 1,164 (2.6%) | <0.001 |
| Metformine | 139 (5.0%) | 527 (1.2%) | <0.001 |
| Warfarin | 96 (3.5%) | 378 (0.8%) | <0.001 |
| Sulfonylureas | 88 (3.2%) | 260 (0.6%) | <0.001 |
| Iron therapy | 156 (5.6%) | 1,566 (3.5%) | <0.001 |
| Betablocker | 828 (29.9%) | 5,182 (11.5%) | <0.001 |
| ACE-inhibitor | 1,066 (38.5%) | 5,555 (12.4%) | <0.001 |
| ARB | 460 (16.6%) | 2,012 (4.5%) | <0.001 |
| Loop diuretics | 374 (13.5%) | 827 (1.8%) | <0.001 |
| Ca-channel blocker | 746 (26.9%) | 4,236 (9.4%) | <0.001 |
| Aspirin | 1,013 (36.6%) | 6,980 (15.5%) | <0.001 |
| Thiazide diuretics | 642 (23.2%) | 3,812 (8.5%) | <0.001 |

**Supplemental Figures**

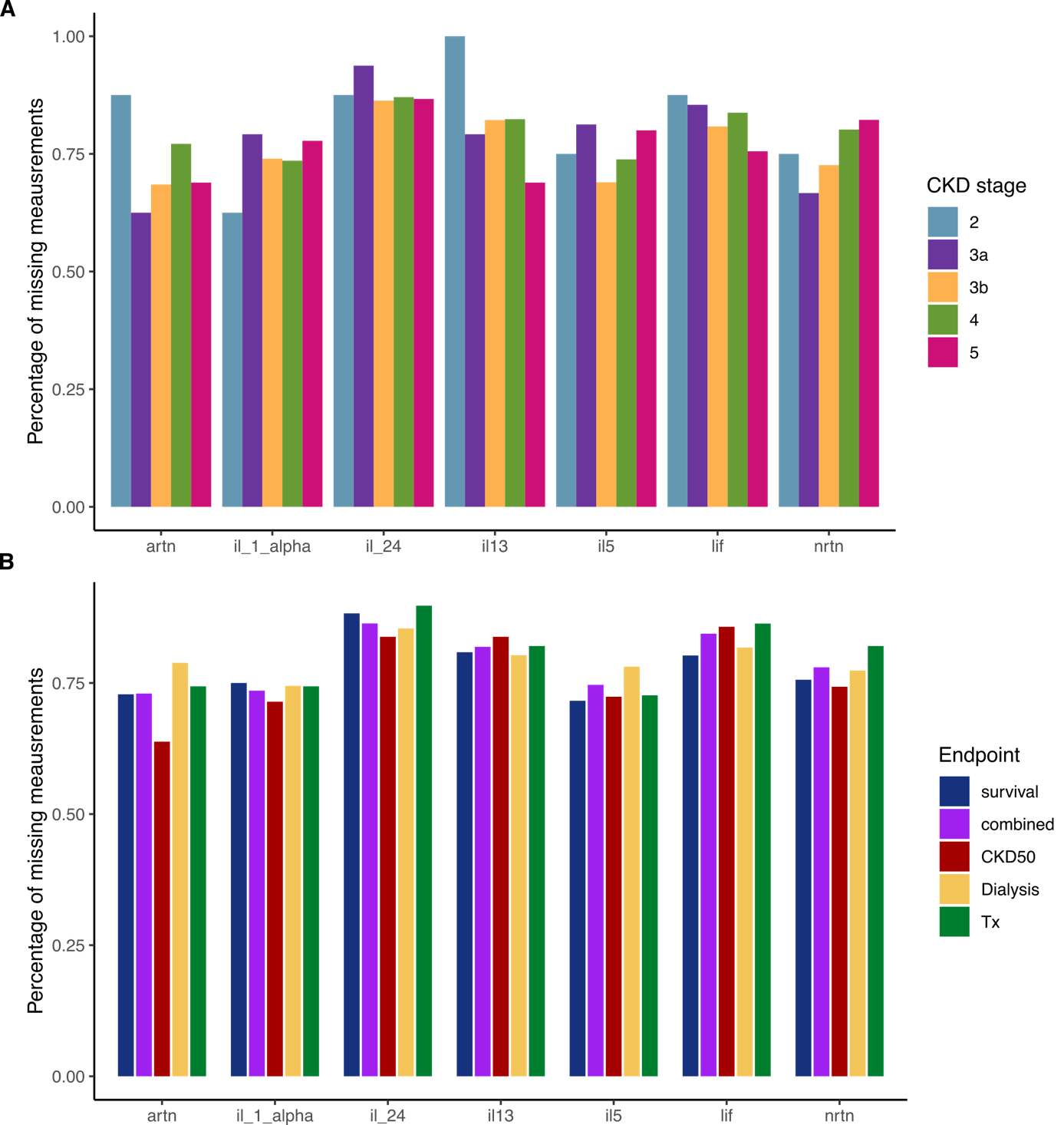

**Figure S1. OLINK features excluded due to missingness in the cohort.** A) Missingness according to CKD stage did not show any patterns. B) Missingness according to endpoint reach similarly shows no pattern.

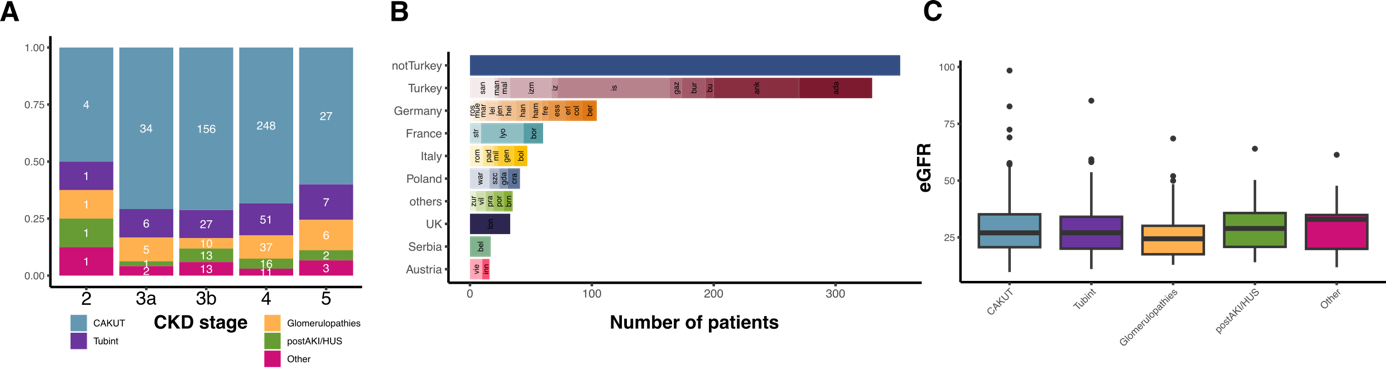

**Figure S2. Baseline characteristics of patients included in this study.** A) Underlying disease according to CKD stage. B) Number of patients recruited per country and study center. ada, Adana, Turkey (Cukurova Universitesi); ank, Ankara, Turkey (Hacettepe Medical Faculty); ank1, Ankara, Turkey (Gazi University Hospital); ank2, Ankara, Turkey (Ankara University Faculty of Medicine); ank3, Ankara, Turkey (Baskent University Faculty of Medicine); ank4, Ankara, Turkey (Diskapi Childrens Hospital); ank5, Ankara, Turkey (Sami Ulus Childrens Hospital); bel, Belgrade, Serbia (University Children’s Hospital); ber, Berlin, Germany (Charité Children’s Hospital); bol, Bologna, Italy (S. Orsola-Malpighi Hospital, Department of Pediatrics); bor, Bordeaux, France (SORARE, Service de Pédiatrie); brn, Bern, Switzerland (Inselspital); bu2, Bursa, Turkey (Dortcelik Children’s Hospital); bur, Bursa, Turkey (Uludag University); col, Cologne, Germany (University Children’s Hospital); cra, Krakow, Poland (University Children’s Hospital); erl, Erlangen, Germany (University Children’s Hospital); ess, Essen, Germany (University Children’s Hospital); fre, Freiburg, Germany (Center for Pediatrics and Adolescent Medicine); gaz, Gaziantep, Turkey; gda, Gdansk, Poland (Medical University); gen, Genova, Italy (Istituto Giannina Gaslini); ham, Hamburg, Germany (UKE University Children’s Hospital); han, Hannover, Germany (Hannover Medical School); hei, Heidelberg, Germany (Center for Pediatrics and Adolescent Medicine); inn, Innsbruck, Austria (Medical University Innsbruck); is1, Istanbul, Turkey (Istanbul Medical Faculty); is2, Istanbul, Turkey (Cerrahpasa Tip Fakultesi); is3, Istanbul, Turkey (Goztepe Educational and Research Hospital); is4, Istanbul, Turkey (Bakirkoy Children Hospital); is5, Istanbul, Turkey (Sisli Educational and Research Hospital); is6, Istanbul, Turkey (Marmara University Medical Faculty); is7, Istanbul, Turkey (Haseki Educational and Research Hospital); izm, Izmir, Turkey (Ege University); iz2, Izmir, Turkey (Tepecik Training and Research Hospital); jen, Jena, Germany (Klinik für Kinder- und Jugendmedizin); lei, Leipzig, Germany (City Hospital St. Georg); lod, Lodz, Poland (Polish Mothers Memorial Hospital Research Institute); lon, London, United Kingdom (Great Ormond Street Hospital); lyo, Lyon, France (Hôpital Femme Mère Enfant & Université de Lyon); mal, Malatya, Turkey (Inonu University Medial School); man, Manisa, Turkey (Celal Bayar University, Pediatric Nephrology Department); mar, Marburg, Germany (KfH Kidney Center for Children); mil, Milano, Italy (Fondazione OSP Maggiore Policlinico); mue, Münster, Germany (University Children’s Hospital); pad, Padova, Italy (University of Padova); por, Porto, Portugal (Hospital Sao Joao); pra, Prague, Czech Republic (University Hospital Motol); rom, Rome, Italy (Bambino Gesu); ros, Rostock, Germany (Children’s Hospital); san, Sanliurfa, Turkey (Sanliurfa Children’s Hospital); str, Strasbourg, France (Hôpital de Hautepierre); szc, Szczecin, Poland (Clinic of Pediatrics); vie, Vienna, Austria (University Children’s Hospital); vil, Vilnius, Lithuania (Vilnius University Children’s Hospital); war, Warsaw, Poland (Children’s Memorial Health Institute); zab, Zabrze, Poland; zur, Zürich, Switzerland (University Children’s Hospital). C) eGFR according to underlying diseases.

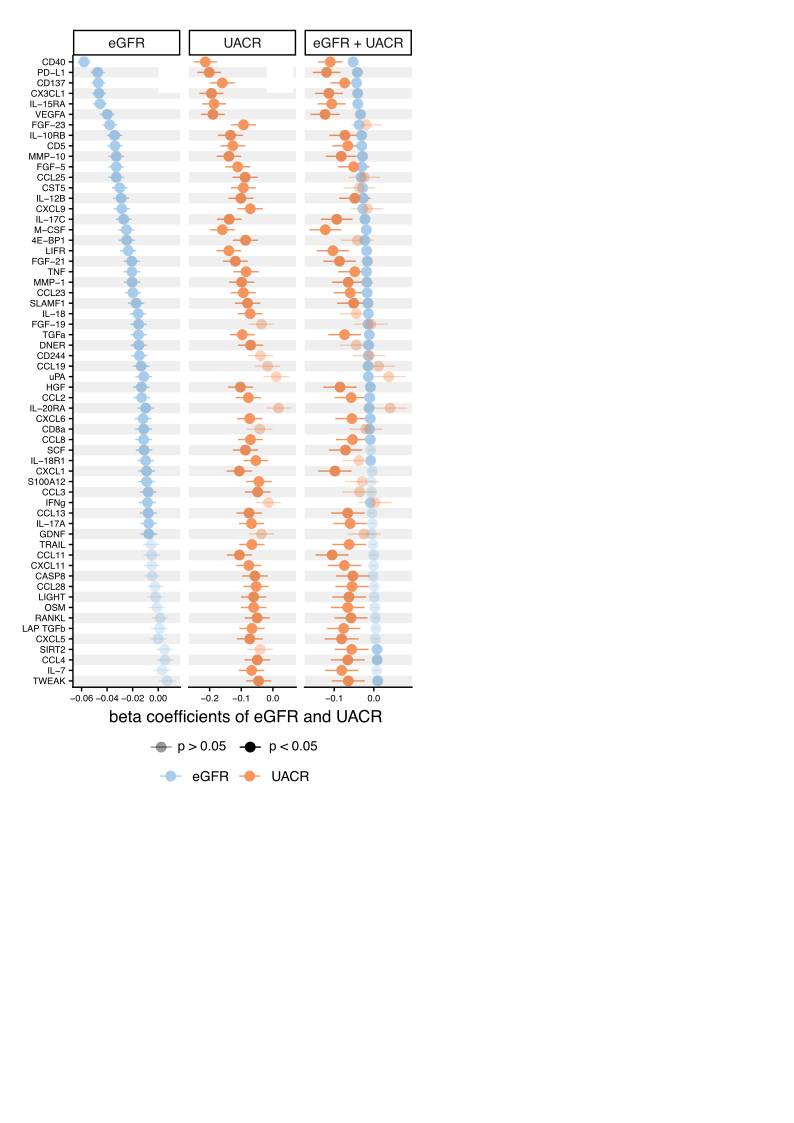

**Figure S3. Beta coefficients of eGFR and UACR for protein levels.** Beta estimates and 95% confidence intervals from linear models with only eGFR (left), urinary albumin to creatinine ratio (UACR, middle) and in a combined model with both (eGFR in blue, UACR in orange, right). FDR adjustment was performed according to Benjamini-Hochberg across models, non-significant associations are shown with reduced opacity.

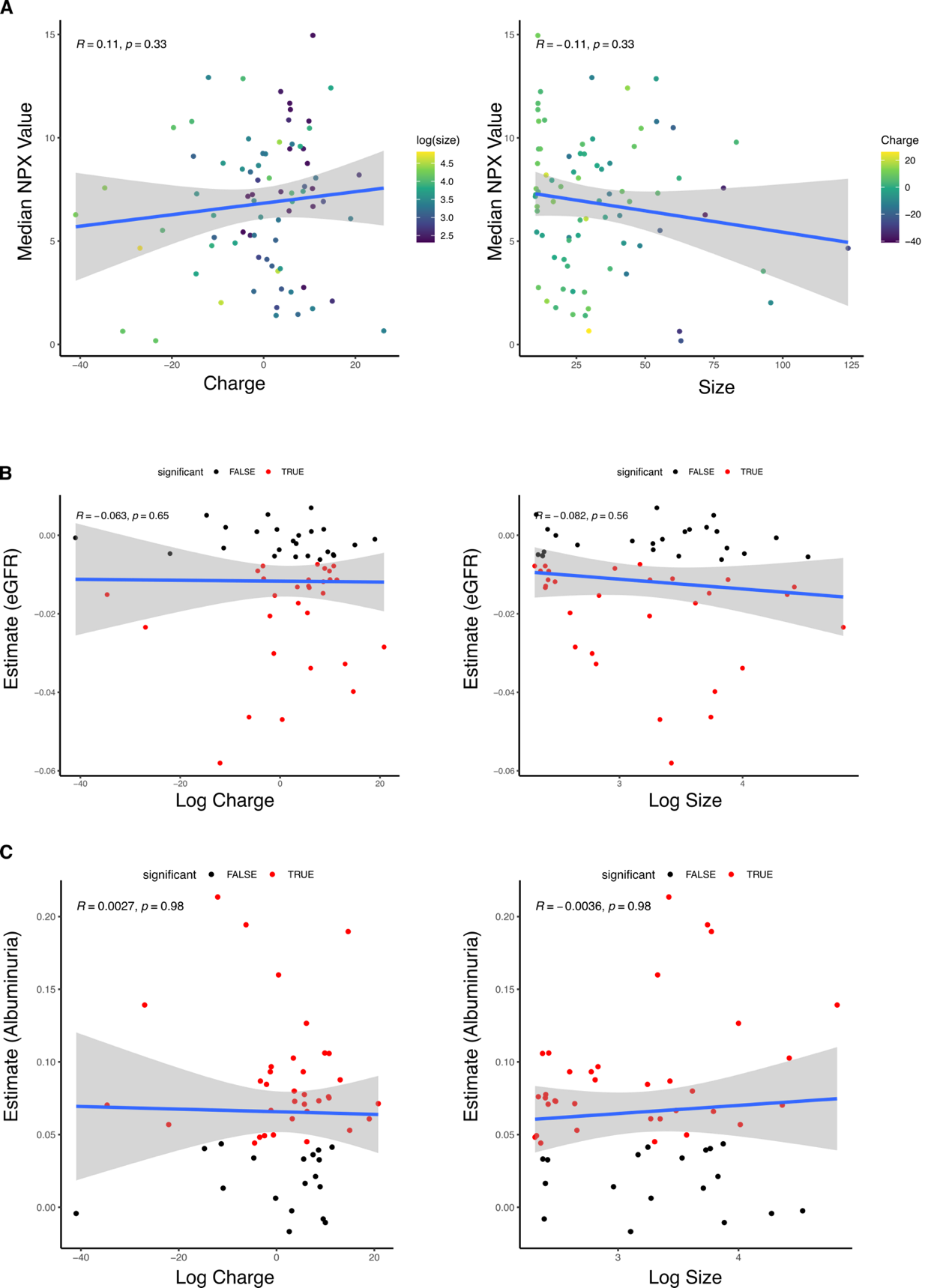

**Figure S4. Association of protein charge and size with the effects of chronic kidney disease.**

A) Correlation of protein charge (left) and size (right) with their median abundance in the 4C cohort. B) Correlation of protein charge (left) and size (right) with the effects of eGFR (beta estimates) on proteins. Color indicates significance after adjustment for multiple testing using linear models (Figure S3). B) Correlation of protein charge (left) and size (right) with the effects of albuminuria (beta estimates) on proteins. Color indicates significance after adjustment for multiple testing using linear models (Fig. S3).

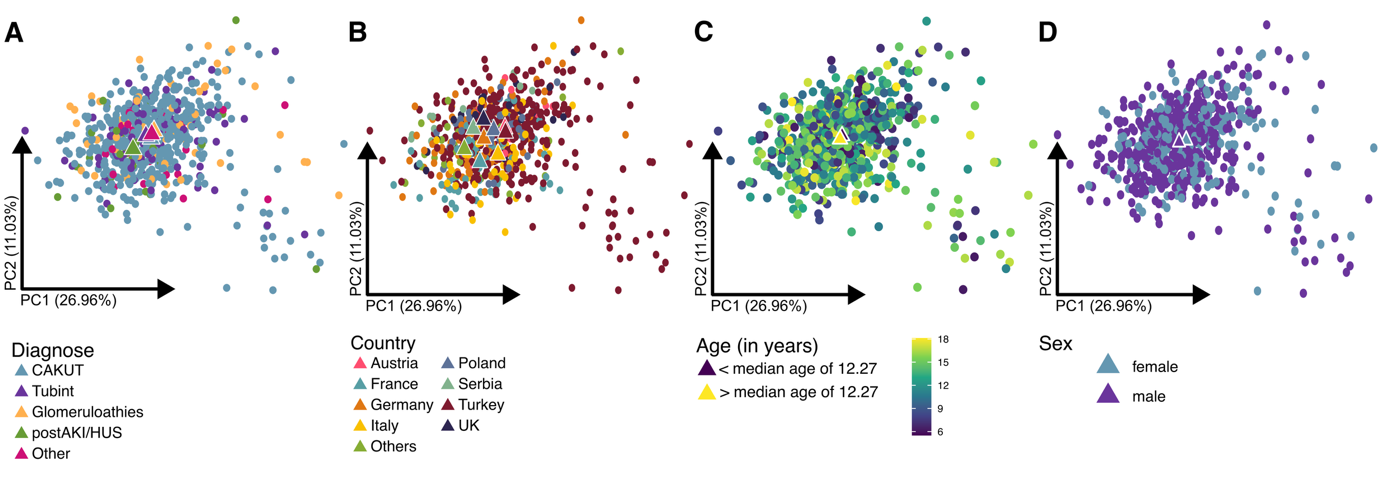

**Figure S5. Principal component analysis (PCA) of inflammation-related plasma proteins**. Colored by A) diagnose, B) country of study center, C) age, and D) sex.

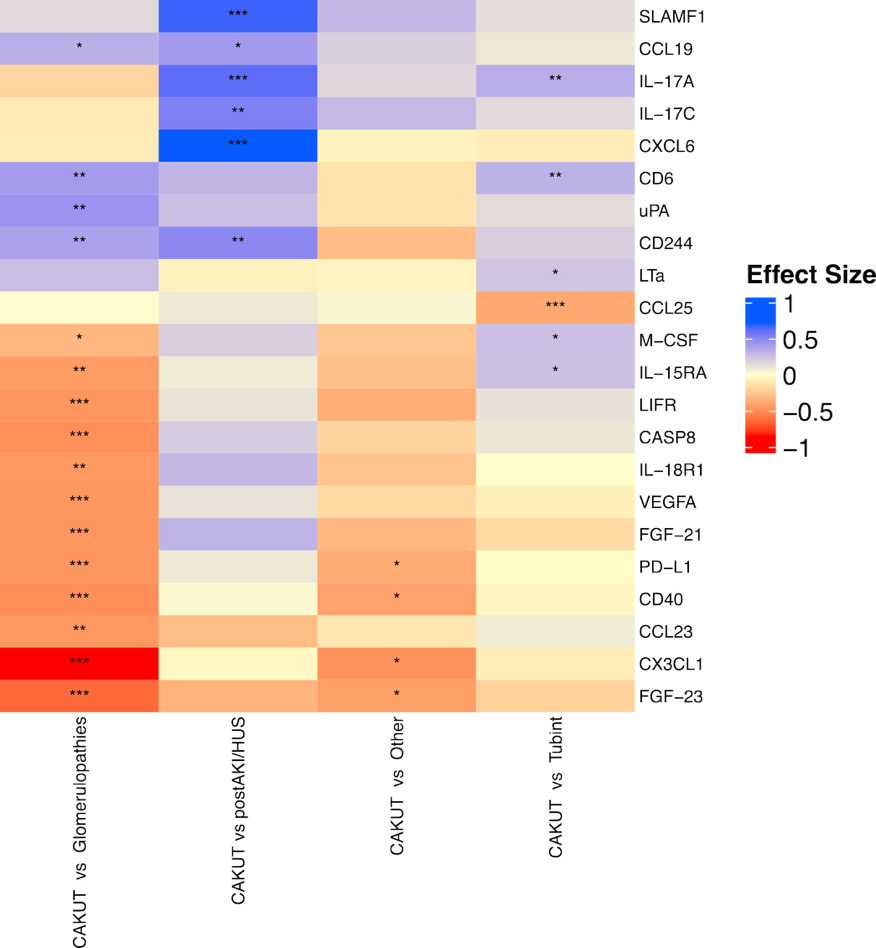

**Figure S6. Effects of underlying kidney disease.** Contrast analysis for underlying kidney diseases versus CAKUT, as largest group of patients**.**

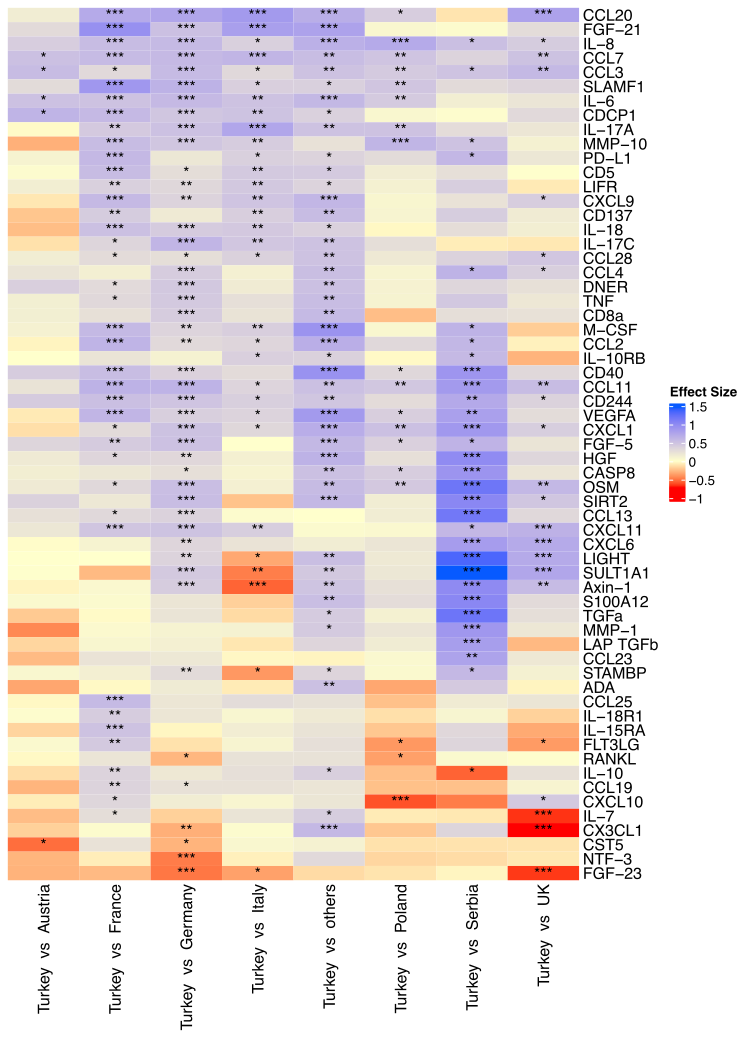

**Figure S7. Effects of country of study center.** Contrast analysis for country of recruitment versus Turkey, with largest group of patients**.**

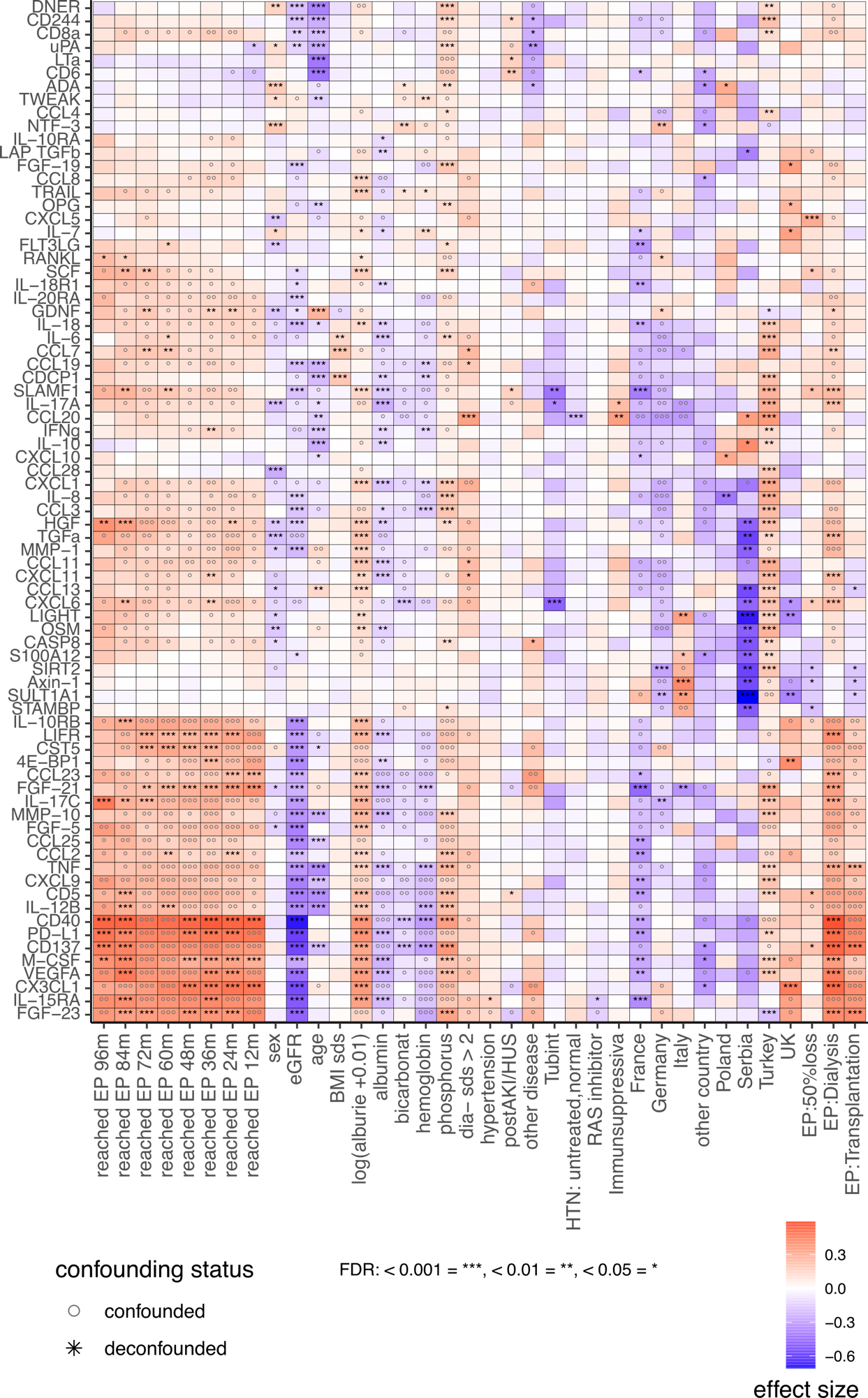

**Figure S8. Confounder-aware statistical modeling of combined kidney endpoint (EP) at discrete time points.** Confounder-aware associations between baseline plasma proteins and the composite kidney endpoint (CKE) at discrete follow-up time points and baseline metavariables. Colors indicate effect size. Symbols denote whether associations remained significant after accounting for potential confounders. Full heatmap corresponding to the subset shown in Fig. 2.

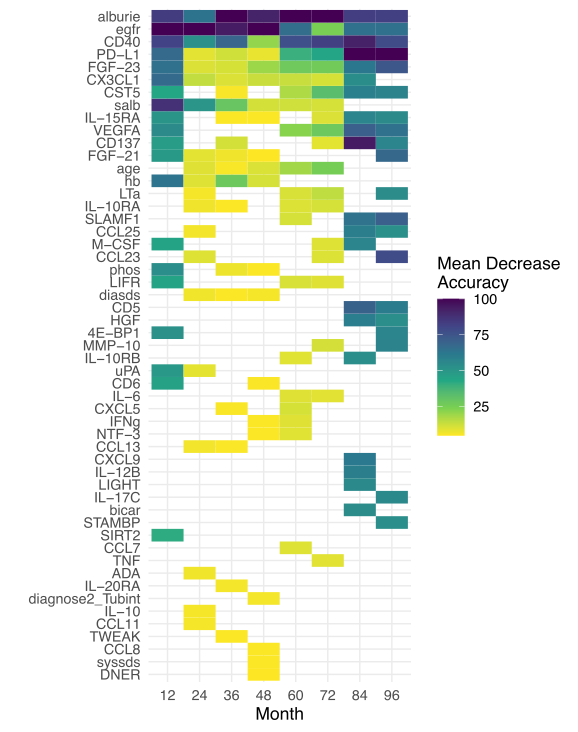

**Figure S9. Variable importance for the random forest classifier of the combined kidney endpoint at discrete follow-up time-points.** Variable importance across combined random forest models for prediction of the CKE at different time points. Colors indicate mean decrease in accuracy, with higher values reflecting greater importance for model performance. Full heatmap corresponding to the subset shown in Fig. 2.

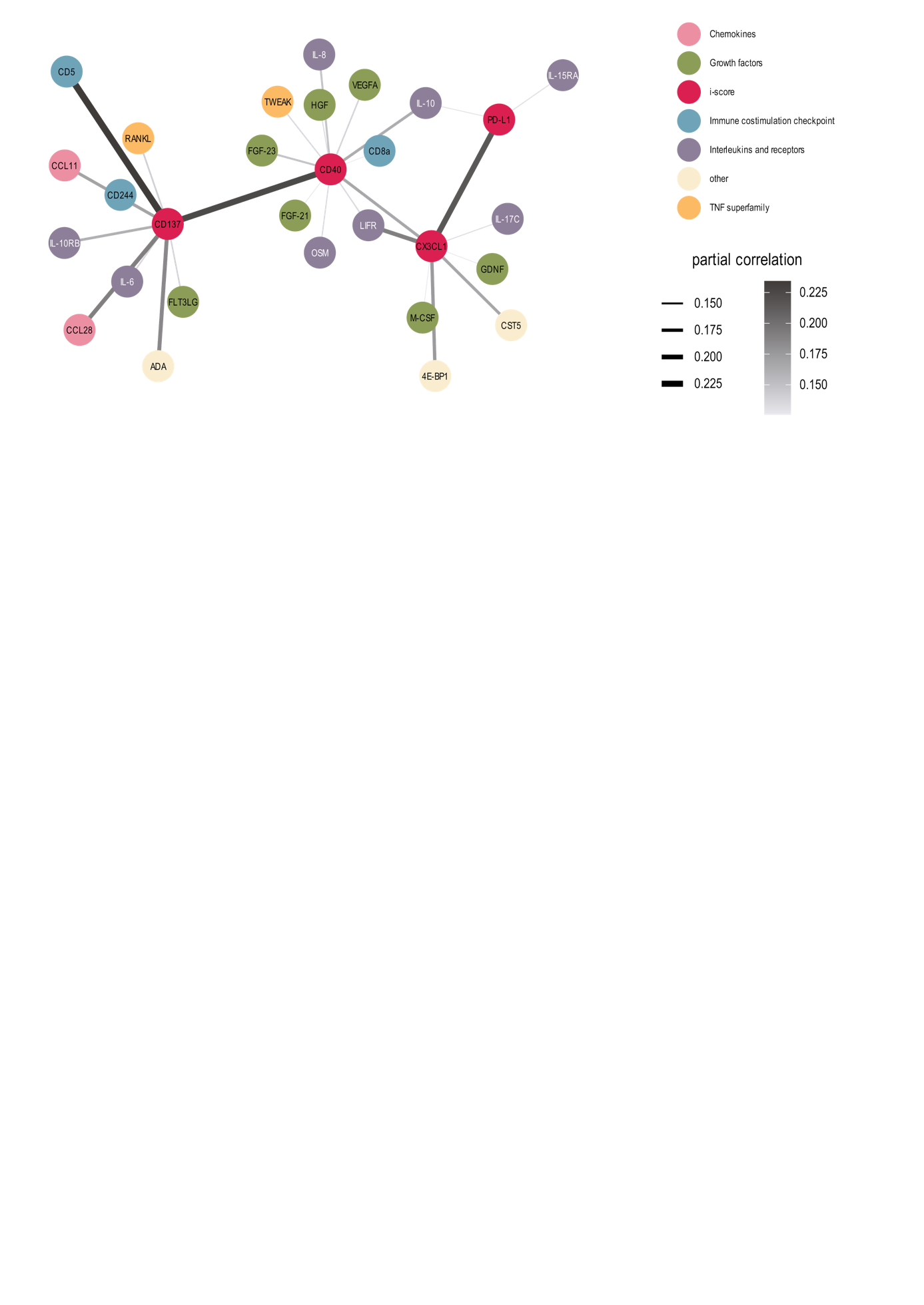

**Figure S10. Partial correlation network of CD40, CD137, CX3CL1 and PD-L1 with other proteins.** Correlations shown are adjusted for eGFR and UACR. Proteins are colored according to biological function. Edge thickness and darker edges represent stronger partial correlation.

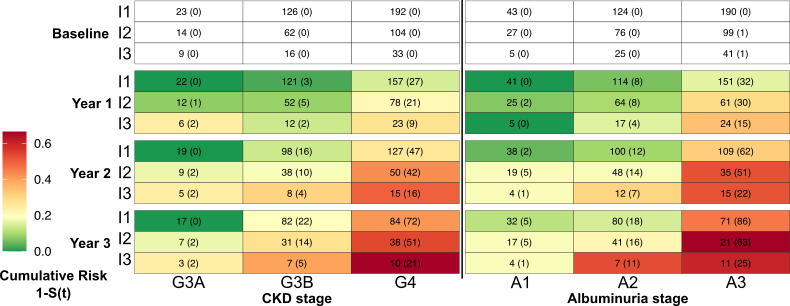

**Figure S11 Cumulative risk for kidney endpoint.** Heatmap showing cumulative risk of the composite kidney endpoint at years 1, 2, and 3 across CKD stage and albuminuria stage, further stratified by iScore category. Number of patients at risk are indicated in the respective boxes and patients reaching the endpoint are shown in brackets.

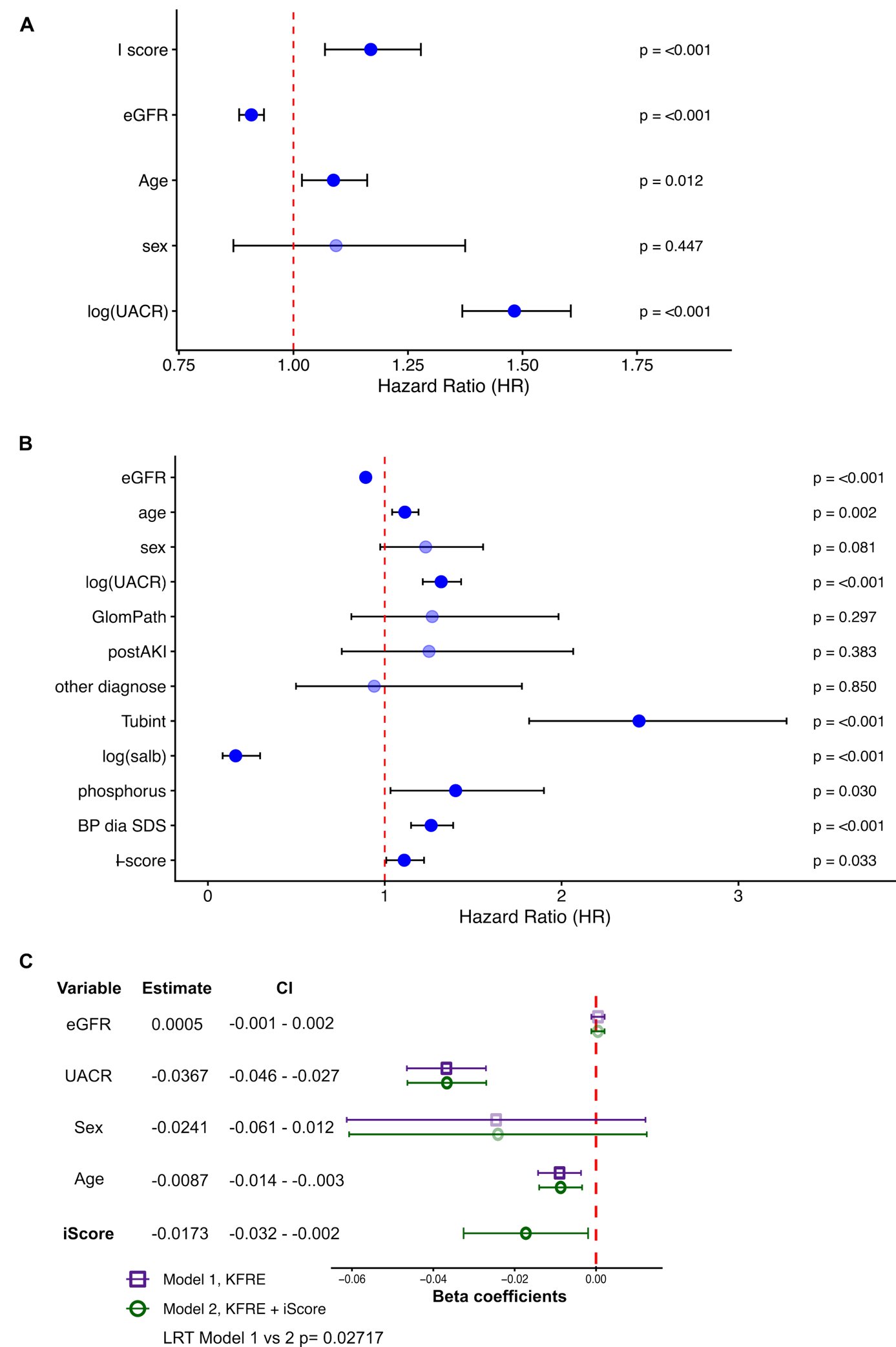

**Figure S12. Cox models for the iScore in the 4C cohort.** Hazard rations (HR) and 95% confidence intervals of all parameters in different Cox models. A) Cox model with iScore KFRE variables. B) Fully adjusted cox model with iScore. (C) Association of iScore with relative eGFR change within the first year after study entry. Linear regression coefficients with 95% confidence intervals are shown for a model with only KFRE variables and KFRE vairables plus iScore. Likelyhood ratio test (LRT) comparing model 1 and model 2 indicates a significantly improved fit through the addition of iScore to KFRE.

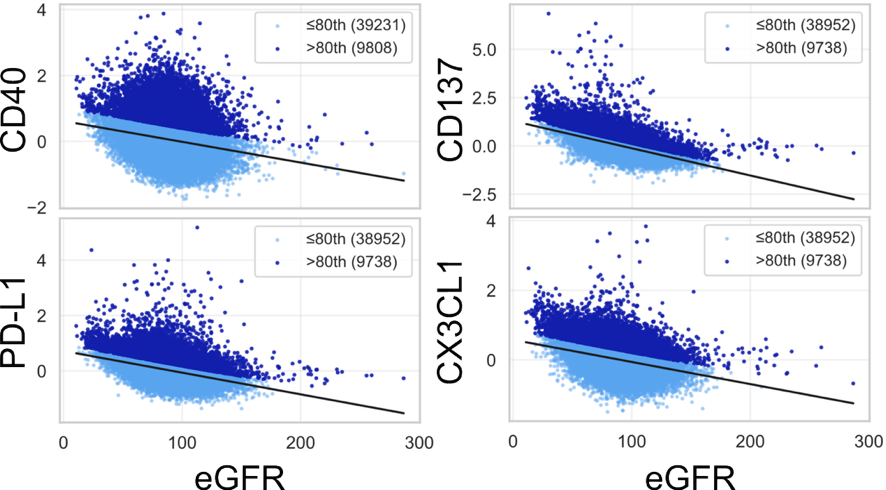

**Figure S13. UKBB iScore residuals.** Baseline levels of CD40, CD137, CX3CL1, and PD-L1 in relation to eGFR. Elevated protein levels were defined as residual values above the 80th percentile after adjustment for eGFR.
